# Insights from Wastewater Surveillance of SARS-CoV-2 in Skilled Nursing Facilities: Comparing Virus Concentration Methods for Wastewater and Correlating Wastewater Virus Concentrations with Clinical Infections, Georgia, USA, 2022

**DOI:** 10.64898/2026.06.01.26354622

**Authors:** Florence Whitehill, Amanda K. Lyons, Bethelhem Abera, Colin Adler, Maria Burgos-Garay, Mariya Campbell, Ariel J. Santiago, Christine Ganim, Jamari Moore, Yimu Cahela, Susanna Lenz, Paige Gable, Magdalena Medrzycki, Maroya Spalding Walters, Amelia Keaton, Peter W. Cook, Yan Li, Ying Tao, Jing Zhang, Lakshmi Malapati, Adam C. Retchless, Suxiang Tong, Margaret Williams, Rodney Donlan, Angela Coulliette-Salmond

**Affiliations:** Division of Healthcare Quality Promotion, National Center for Emerging and Zoonotic Infectious Diseases, Centers for Disease Control and Prevention, 1600 Clifton Rd., Atlanta, GA 30329, USA; Coronavirus and Other Respiratory Viruses Division, National Center for Immunization and Respiratory Diseases, Centers for Disease Control and Prevention, 1600 Clifton Rd., Atlanta, GA 30329, USA; United States Public Health Service, 1101 Wootton Pkwy., Plaza Level, Rockville, MD 20853, USA

**Keywords:** SARS-CoV-2, skilled nursing facilities, wastewater surveillance, virus concentration

## Abstract

To understand the utility of healthcare facility-level wastewater surveillance (WWS) for severe acute respiratory syndrome coronavirus 2 (SARS-CoV-2), it is important to correlate wastewater SARS-CoV-2 RNA detection with the number of clinical infections. WWS for SARS-CoV-2 was performed at three skilled nursing facilities (SNFs) over 25 weeks. Electronegative membrane filtration (enMF) and Nanotrap® Magnetic Virus Particles (NP) virus concentration methods were compared. Extracts were tested by droplet digital polymerase chain reaction. Spearman’s correlations (ρ) between wastewater virus RNA concentrations and infection counts were calculated.

From split wastewater samples, enMF recovered higher SARS-CoV-2 RNA concentrations than NP. Combining data from all facilities, the median concentrations were 53.0 versus 38.6 gc/100 mL for enMF and NP, respectively (p=0.001). Using enMF, correlations were moderate to strong at SNF A (ρ ranged 0.67 to 0.86, all p-values <0.001). Weak to moderate correlations can be explained by the sampled manhole not representing the entire facility (SNF B, ρ ranged 0.47 to 0.72, p-values ranged <0.001 to 0.12) and longitudinal data gaps from summer heat and equipment maintenance (SNF C, ρ ranged 0.14 to 0.59, p-values ranged 0.52 to <0.01). WWS can be a valuable tool for tracking dynamics of SARS-CoV-2 infections in healthcare facilities.

**Highlights:**

1. Healthcare facility-level wastewater surveillance may be useful as a complementary surveillance method for infectious disease targets, including SARS-CoV-2.
2. Two virus concentration methods were compared, electronegative membrane filtration (enMF) and Nanotrap® Magnetic Virus Particles (NP).
3. Spearman’s correlations (ρ) between wastewater virus RNA concentrations and infection counts were calculated among several subpopulations at three skilled nursing facilities.

**Graphical Abstract:** 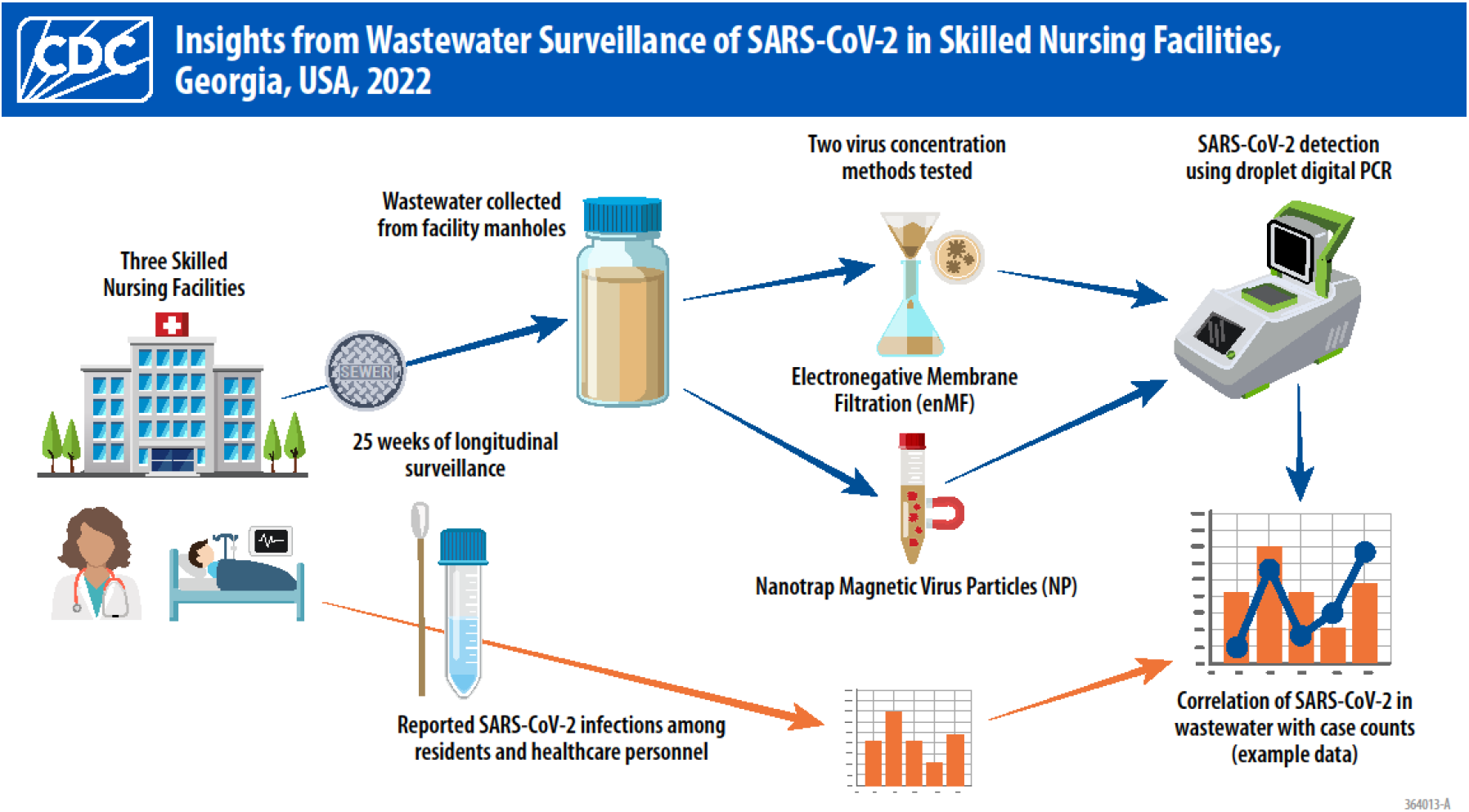

## Introduction

Older adults are disproportionally affected by severe acute respiratory syndrome coronavirus 2 (SARS-CoV-2) infection, having both higher morbidity and mortality rates (1, 2). The COVID-19 pandemic also disproportionately affected residents in long-term care facilities such as skilled nursing facilities (SNFs), whose populations have high proportions of people ≥ 65 years and who require specialized medical care and assistance with daily living activities (3, 4). Syndromic surveillance for SARS-CoV-2 infections can be time- and resource-intensive, and relies on the recognition of symptomatic cases, potentially missing cases who are asymptomatic or not yet symptomatic (5-7). Wastewater surveillance (WWS) at the healthcare facility-level (hereafter “facility level”) may be useful as a complementary surveillance method for enhanced detection of concerning infectious disease targets, including SARS-CoV-2. While WWS has been used widely for many applications at both the community and facility levels, it is important to determine the utility of facility-level WWS for infectious disease targets before more widespread use can be efficiently implemented. Likely benefits of facility-level WWS include early detection of outbreaks and improved surveillance of healthcare–associated infections, which could potentially lead to life and cost savings.

This study contributes to the growing scientific understanding of the feasibility and operability of facility-level WWS for SARS-CoV-2 (8-10). The research team previously completed a pilot study at one SNF in the metropolitan Atlanta area to assess the feasibility of detecting SARS-CoV-2, endogenous controls, and antimicrobial resistance targets from wastewater samples (11). The study presented here is a continuation of this work focused on longitudinal measurements needed to correlate SARS-CoV-2 detection in wastewater with SARS-CoV-2 infections at three SNFs in Georgia. We also present a comparison of two virus concentration methods, electronegative membrane filtration (enMF) and Nanotrap® Magnetic Virus Particles (NP).

## Materials and Methods

This study was approved by the CDC COVID-19 Response Laboratory Task Force and met the CDC safety policies and recommendations in the CDC/NIH Biosafety in Microbiological and Biomedical Laboratories (6). This activity was reviewed by CDC and was conducted consistent with applicable federal law and CDC policy^§^.

This study met the minimum information for publication of quantitative digital PCR experiments (dMIQE). Details are available in Supplement 3 (12).

### Facility Selection and Epidemiologic Data Collection

#### Facility Selection

Beginning in 2021, the research team consulted the Georgia Department of Public Health (GDPH) Healthcare Associated Infections Team to identify SNFs in the Atlanta metropolitan area that were amenable to WWS participation. The research team and GDPH considered a facility’s COVID-19 case counts in 2021, the strength of a facility’s relationship with GDPH, and the facility’s capacity to participate in WWS before reaching out to gauge interest in participation. In total, 18 facilities were contacted. Seven facilities responded with interest in participating.

An enrollment questionnaire was administered at seven SNFs during initial onsite visits (Supplement 1). The research team filled out the questionnaire through field site observations and asking questions of the facility administrators, along with the Infection Preventionist or Director of Nursing. This questionnaire collected information about wastewater access points, types of healthcare provided, current resident census, COVID-19 vaccination rates among residents and healthcare personnel, COVID-19 screening practices, fecal waste management, environmental cleaning practices, and building layout. Three SNFs, designated as SNFs A, B and C, were recruited to participate based upon the following criteria: 1) resident census ≥ 75, 2) within a 90-minute drive of CDC campus, and 3) safely accessible manhole or wastewater access point. SNF A participated in longitudinal wastewater and epidemiologic data collection from February through July 2022 (25 weeks, n= 25 wastewater samples). SNF B participated from March through September 2022 (27 weeks, n=25 wastewater samples). SNF C participated from June through December 2022 (29 weeks, n=25 wastewater samples). Of note, SNF A participated in the previous eight-week pilot study with the research team to determine the feasibility of facility-level WWS and the detection of SARS-CoV-2 in wastewater (11).

#### Epidemiologic Data Collection

For statistical analyses, infections per week was defined as the number of residents or healthcare personnel with new laboratory-confirmed SARS-CoV-2 infection by antigen or polymerase chain reaction (PCR) test in a given week, excluding those who had a negative PCR test (i.e., referred to as a nucleic acid amplification test (NAAT) in public health laboratories) after an antigen test as defined by the National Healthcare Safety Network (NHSN) Long-Term Care Facility COVID-19 Report (13).

One resident subpopulation, residents contributing fecal matter to wastewater, was defined as residents who either contributed fecal matter to the wastewater through direct toilet usage (toileting) or contributed by having their feces disposed of in the toilet by healthcare personnel after being collected, such as in a diaper or bedside commode. Among this subpopulation we defined infections per week as the total number of laboratory-confirmed SARS-CoV-2 infections in the facility each week.

Additional epidemiologic data were acquired from the facility’s healthcare team which typically included the Director of Nursing and the Infection Preventionist. The facility’s healthcare team completed and returned a standardized questionnaire each calendar week that wastewater sampling occurred. This questionnaire collected information about COVID-19 outbreaks at the facility, SARS-CoV-2 infections among a subpopulation of residents who contributed fecal matter to the wastewater, vaccination rates among all residents and healthcare personnel, number of visitors, and any chemical disinfectants disposed of down the drain (i.e., added to the wastewater) (Supplement 1). The completed questionnaires were emailed to the research team who entered the data into a Research Electronic Data Capture (REDCap) database (14, 15).

#### Plumbing Tracer Analysis and SNF B Data Re-collection

A visual and viral tracers analysis, described in Coulliette-Salmond et al., was conducted at each facility to confirm wastewater effluent was collected from the entire resident population (16). At SNF B, the plumbing tracer study revealed the manhole used for longitudinal wastewater collection only collected effluent from one wing of the building. The research team contacted SNF B to re-collect epidemiologic survey data specific to the one wing of the facility from which effluent was sampled using an abbreviated retrospective data collection tool (Supplement 1). Retrospective data collection at SNF B was performed by extracting data from notes by the Director of Nursing and limited chart review due to time constraints.

### Wastewater Methods

#### Wastewater Sampling

Wastewater effluent was collected weekly at each facility during the study period. Wastewater was accessed at manholes on the facilities’ properties. A 24-hour composite sample was collected at each sampling event using an Avalanche Portable Refrigerated Autosampler (Teledyne ISCO, Lincoln, NE, USA). Samples were transported on ice in a cooler to the CDC laboratory for same-day processing. See Supplement 2 for results of the measured wastewater quality parameters, which included temperature (°C), pH, electrical conductivity (µS/cm), total dissolved solids (mg/mL), total suspended solids (mg/mL), biochemical oxygen demand (mg/mL), total organic carbon (mg/mL), total organic nitrogen mg/mL), total coliforms (log_10_ MPN/100 mL), and Escherichia coli (log_10_ MPN/100 mL).

#### Virus Concentration and Extraction

Wastewater samples were split and concentrated using two separate methods for comparison: electronegative membrane filtration (enMF) and Nanotrap® Magnetic Virus Particles (NPs). Details of these procedures are described in Santiago et al. with slight modifications: when homogenizing enMFs, 0.5 mm glass, PowerBead tubes (Qiagen, Germantown, MD USA) were substituted for the 0.70 mm garnet PowerBead tubes (discontinued by manufacturer); incubation of NPs in wastewater, as well as removal of NPs by magnetic rack was reduced to 10 min each; recovered NPs were re-suspended in nuclease-free water (11). Viral RNA was extracted using the AllPrep® PowerViral® DNA/RNA Kit (Qiagen, Germantown, MD USA) according to manufacturer’s instructions for RNA isolation, as previously described in Santiago et al. (11).

#### Detection and Quantification of SARS-CoV-2 and Controls

Droplet digital PCR (ddPCR) assays for the detection and quantification of SARS-CoV-2, its viral surrogates (Bovine Respiratory Syncytial Virus [BRSV], human coronavirus OC43 [HCoV-OC43]), and the endogenous control (prototypic crAssphage *Carjivirus communis*) were performed using a QX200 automatic droplet generator system (Bio-Rad, Hercules, CA, USA). RNA extracts were used as template and analyzed using One-Step RT-ddPCR Advanced Kit for Probes (Bio-Rad, Hercules, CA, USA) or ddPCR Supermix for Probes kit (No dUTP; Bio-Rad, Hercules, CA, USA). SARS-CoV-2 and BRSV were detected using the 2019-nCoV CDC triplex assay (dEXS28563542, Bio-Rad, Hercules, CA, USA), where primers and probes were purchased from IDT (Coralville, IA, USA). HCoV-OC43 and *C. communis* were each detected using a singleplex assay. For the triplex assay, 8 µL of RNA was added to 5.5 µL of One-Step Advanced Supermix, 1.1 µL of 300 mM dithiothreitol (DTT), 2.2 µL of reverse transcriptase (RT), 2.2 µL of 10X triplex probe assay, and 3.0 µL RNase-free water in a final volume of 22 µL. For the singleplex assay used to detect HCoV-OC43, 8 µL of RNA was added to 5.5 µL of One-Step Advanced Supermix, 1.1 µL of 300 mM dithiothreitol (DTT), 2.2 µL of RT, 1 µL of forward primer, 1 µL of reverse primer, 0.3 µL of probe (final concentrations were 900 nM of primers and 250 nM of probe), and 2.9 µL RNase-free water in a final volume of 22 µL. For the singleplex assay used to detect *C. communis*, 2 µL of DNA was added to 11 µL of ddPCR Supermix for Probes (No dUTP), 1 µL of forward primer and 1 µL of reverse primer, 0.3 µL of probe (final concentrations were 900 nM of primers and 250 nM of probe), and 6.7 µL nuclease-free water in a final volume of 22 µL. Sequences for all primers and probes are detailed in Table S1B in Santiago et al. (11). For droplet generation, 20 µL of reaction mixture was added to each well of a semi-skirted, 96-well ddPCR plate and loaded into the automatic droplet generator. Plates were then sealed using the PX1 Plate Sealer and transferred to a C1000 touch thermal cycler (Bio-Rad, Hercules, CA). The amplification of the triplex assay and singleplex (HCoV-OC43) were performed as follows: 1 cycle of 60 min at 50°C and 10 min at 95°C, 40 cycles of 30 s at 94°C and 1 min at 55°C, 1 cycle of 10 min at 98°C and a 30 min hold at 4°C. The amplification of the singleplex assay for *C. communis* was performed as follows: 1 cycle of 10 min at 95°C, 40 cycles of 30 s at 94°C and 1 min at 60°C, 1 cycle of 10 min at 98°C and a 30 min hold at 4°C. All steps were run using a ramp rate of 2°C/s.

Each run was performed using positive controls (synthetic 2019_nCoV RNA control [CDC], BRSV, HCoV-OC43 and *C. communis*) and negative controls (extraction blanks and non-template controls [NTC]). A “no-reverse transcriptase” control (i.e., No-RT control per dMIQE, Supplement 3) was included to verify that there was no amplification in the absence of reverse transcriptase. Technical triplicates were run for each sample including controls. To test for PCR inhibition, each template was run and analyzed using two dilution factors (undiluted and 1:2 fold) for all assays. After amplification, plates were transferred to the droplet reader. Reactions with less than 10,000 total droplets were repeated. Data were analyzed using QuantaSoft^™^ AnalysisPro Software and reported as genomic copies per microliter (gc/µL) of template. The threshold was manually set by inspecting the 2D plot to clearly separate positive and negative droplet clusters. For analysis, the mean of triplicate samples was determined. Samples were considered positive if ≥ 3 positive droplets were detected for SARS-CoV-2 N1 or N2 markers in at least 2 of 3 triplicate samples (i.e., limit of quantification; Supplement 4 Table S4.1). Results were considered ‘detectable but not quantifiable’ (DNQ) in two scenarios: (1) if there was at least one positive droplet, but less than 3 positive droplets, in any/all of the triplicates; or (2) for samples that had 1 triplicate with > 3 droplets but the associated two replicates were less than 3 droplets. The limit of detection (LOD), which was determined in a separate analysis with spiked samples, was defined as the concentration at which > 60% of replicates were detectable was 0.125 and 0.126 gc/µL template, for N1 and N2 targets, respectively (17). A sample was considered negative if no positive SARS-CoV-2 droplets were detected. Dimensional analysis using Equation 1 produced final results expressed as log_10_ genomic copies (gc)/100 mL in Figures 1 and 2 and otherwise as gc/100 mL (18, 19).

**Equation 1:**

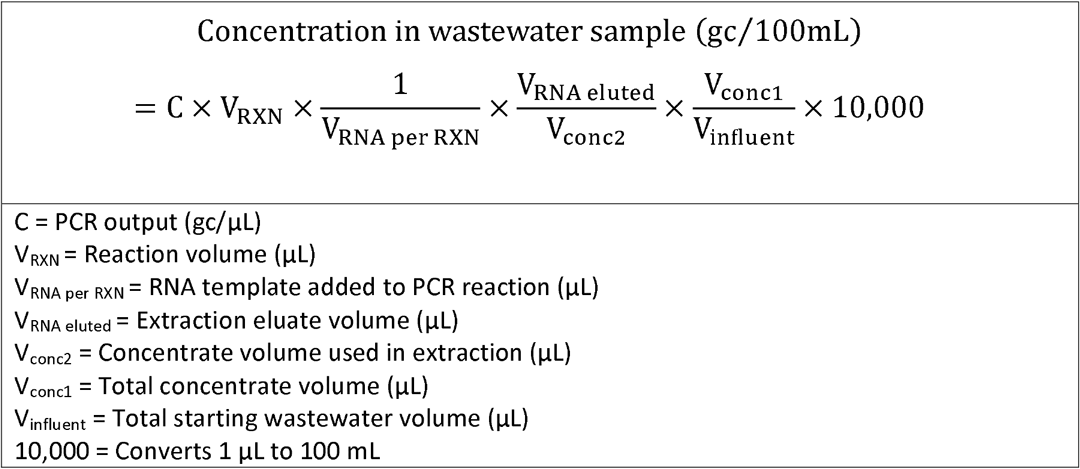

**Figure 1:**
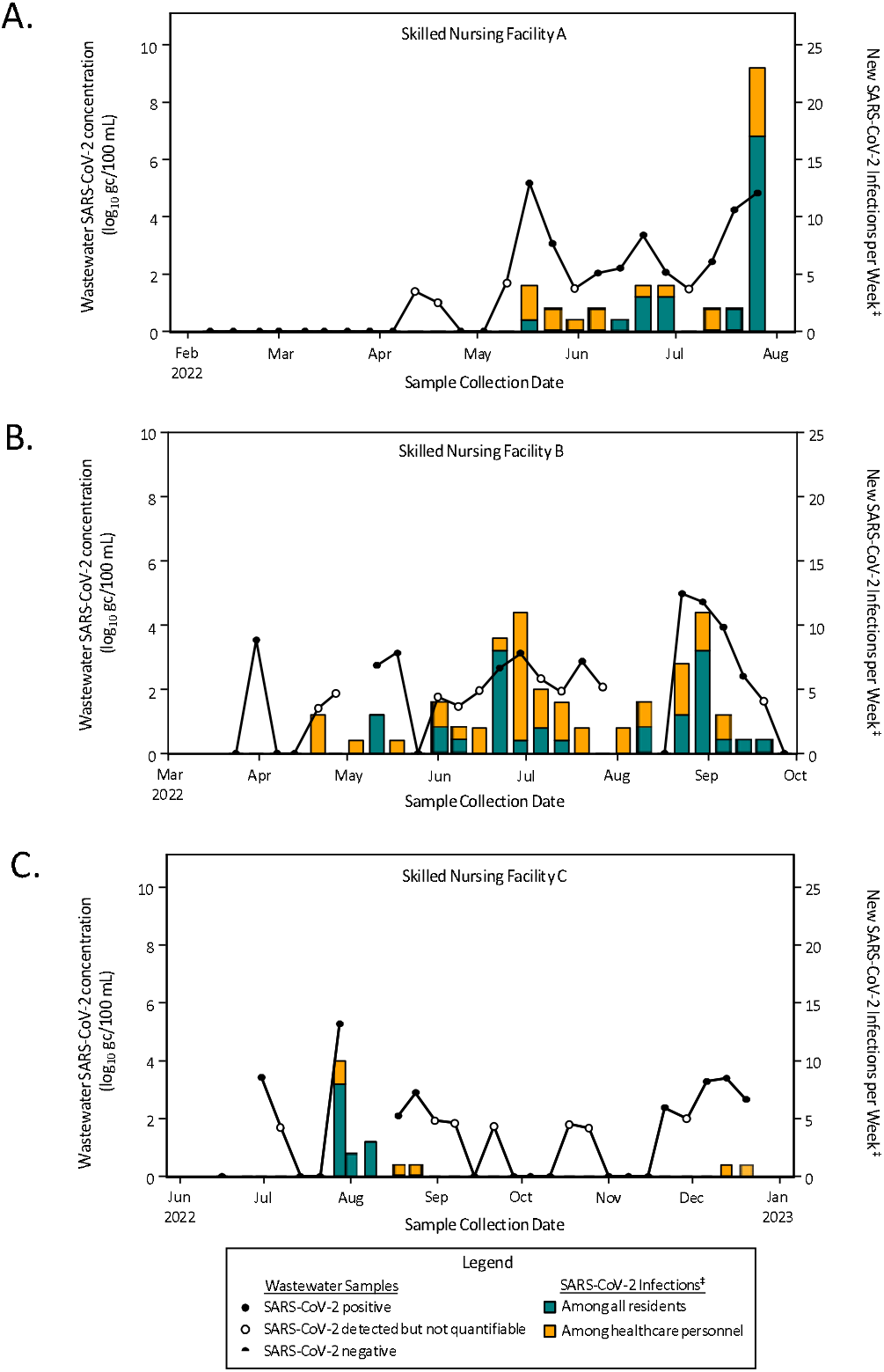
Wastewater SARS-CoV-2 concentration (using the enMF concentration method^†^) and SARS-CoV-2 infection counts among residents and healthcare professionals over 25 weeks at three skilled nursing facilities (panels A – C), Atlanta, GA, 2022 Figure 1 Footnotes: *Abbreviations: SARS-CoV-2, severe acute respiratory syndrome coronavirus 2; gc, genome copies; enMF, electronegative membrane filtration ^†^Results presented in this figure were generated with the enMF virus concentration method. Results generated with the nanoparticle virus concentration method are presented in Supplement 4, Figure S4.1. ^‡^Data sources for SARS-CoV-2 infections: counts among healthcare personnel as well as among all residents at skilled nursing facilities (SNFs) A and C were obtained from the National Healthcare Safety Network Long-Term Care Facility COVID-19 Report; infection counts among residents in one wing of at SNF B were obtained from the condensed epidemiologic data re-collection tool (Supplement 1).

**Figure 2:**
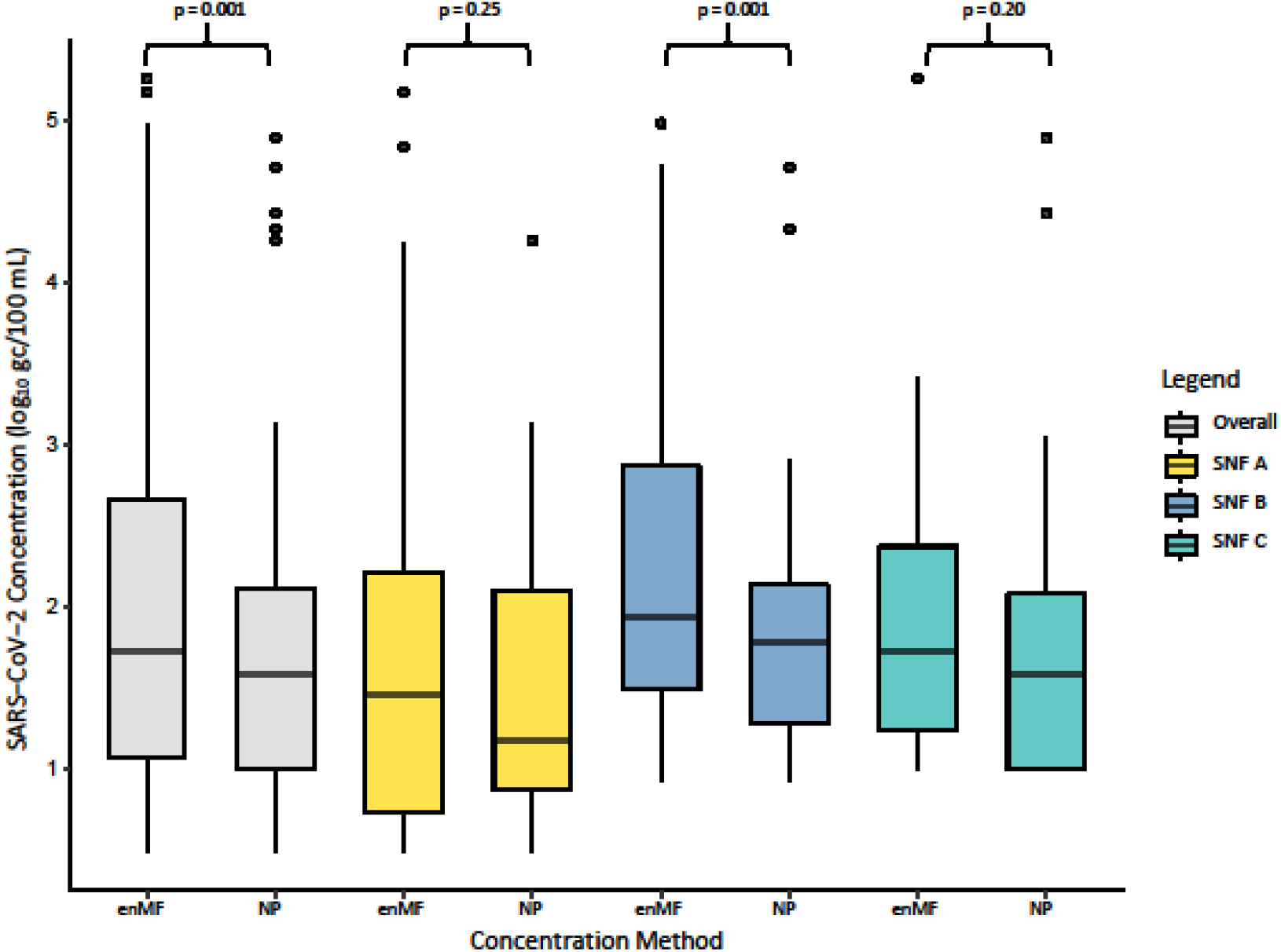
Comparison of SARS-CoV-2 RNA concentration in wastewater using two virus concentration methods, enMF vs. NP, at three skilled nursing facilities (n = 25 samples per facility) Atlanta, GA, 2022 Figure 2 footnotes: Abbreviations: SARS-CoV-2, severe acute respiratory syndrome coronavirus 2; enMF, electronegative membrane filtration; NP, nanoparticle; gc, genome copies; SNF = skilled nursing facility *p-values represent the statistical significance of the difference between the median viral RNA concentrations within each category, α = 0.05

### Statistical Analyses

A wastewater sample was considered positive if N1 or N2 was positive by ddPCR droplet count as described above. This definition was applied consistently across statistical analyses. Prior to statistical analysis, non-detect values were imputed using half the smallest non-zero concentration value. N1 and N2 concentrations were then combined using the geometric mean of the two targets. The distribution of combined N1/N2 results were assessed using the Shapiro-Wilk test of normality, which indicated a non-normal distribution therefore non-parametric tests were used for downstream analysis.

SAS software (version 9.4, 2023, Cary, North Carolina), Microsoft® Excel® (version 2408, Microsoft Corporation, Redmond, Washington), and R (version 4.4.0, R Core Team, Vienna, Austria) were used for the statistical analyses.

#### Comparison of Concentration Methods

Differences in the median SARS-CoV-2 (combined N1/N2) concentration was compared across the enMF and NP concentration methods using the Wilcoxon signed-rank test (α = 0.05).

#### Sensitivity and Specificity

The sensitivity and specificity of using wastewater SARS-CoV-2 (combined N1/N2) concentrations to detect the presence/absence of SARS-CoV-2 infections were calculated for each facility separately and for all three facilities combined. A facility was considered disease positive if, during the week of wastewater collection, it had at least one reported SARS-CoV-2 infection among all residents or among all residents and healthcare personnel combined.

#### Spearman’s Correlations

Spearman’s correlation was used to evaluate the relationship between the SARS-CoV-2 (combined N1/N2) concentration in wastewater and SARS-CoV-2 infections in a facility each week over 25 weeks. Spearman’s correlations were assessed separately for results from the two concentration methods, NP and enMF. The strength and direction of correlations between wastewater SARS-CoV-2 concentration and reported infection counts among several subpopulations in the facilities were determined: all residents, healthcare personnel, all residents and healthcare personnel combined, and residents contributing fecal matter to wastewater. Spearman’s correlation coefficient (ρ) values > 0 and < 0.4 were considered weak, ≥ 0.4 and < 0.7 considered moderate, and ≥ 0.7 and < 1 considered strong (20). Significance level was set at α = 0.05.

### Viral Sequencing

Eighteen of 29 wastewater samples that were positive for SARS-CoV-2 and had an estimated ddPCR gc/µl concentration that equated to a real-time PCR quantification cycle (Cq) < 35 were selected for genomic sequencing across non-standardized time points from SNFs A (n=7), B (n=6), and C (n=5). Real-time PCR was performed on the selected samples to confirm Cq < 35. Two of the 18 samples did not have confirmed real-time PCR Cq < 35 and 5 of 18 samples did not have greater than 60% genome coverage and therefore did not meet the standards for sequencing (21). As a result, eleven of the 18 wastewater extracts were successfully sequenced, available in NCBI BioProject PRJNA1280116. Further details regarding sequencing methods are provided in Supplement 4.

## Results

### Overview of Participating Facilities

The weekly mean resident census was 169.8, 79.7, and 92.4 at SNFs A, B, and C, respectively (Table 1). The weekly mean proportion of residents contributing fecal matter to the wastewater was 46.6%, 81.7%, and 36.1%, at SNFs A, B, and C, respectively. All facilities provided skilled nursing, wound, ostomy, and indwelling medical device care. SNFs B and C also provided inpatient rehabilitation. None of the three facilities provided ventilator care. The mean proportion of all residents who were fully vaccinated against SARS-CoV-2 were 91.6%, 98.3%, and 89.3%, at SNFs A, B, and C, respectively (Supplement 4, Table S4.2). During this study period from February through December 2022, individuals were considered fully vaccinated if they had completed the primary series of COVID-19 vaccines, which included two doses of the Pfizer-BioNTech or Moderna vaccines, or one dose of the Johnson & Johnson vaccine (22). Additional information about vaccination rates among residents and healthcare personnel is provided in Supplement 4, Table S4.2.

**Table 1:**
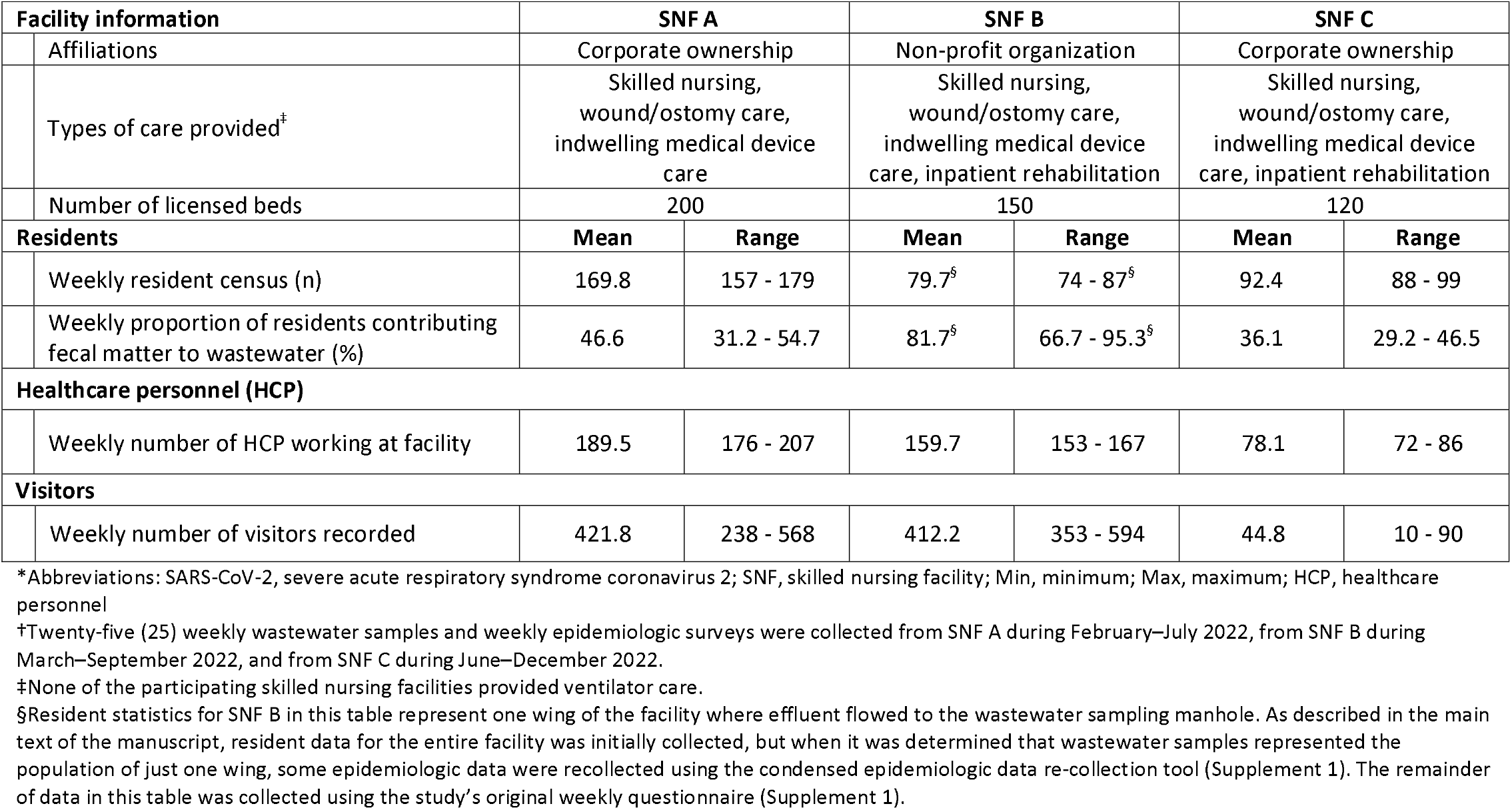
Summary information for three skilled nursing facilities enrolled in longitudinal SARS-CoV-2 wastewater surveillance study, Atlanta, GA, 2022^†^.

| Facility information |  | SNF A |  | SNF B |  | SNF C |  |
| --- | --- | --- | --- | --- | --- | --- | --- |
|  | Affiliations | Corporate ownership |  | Non-profit organization |  | Corporate ownership |  |
|  | Types of care provided<sup>‡</sup> | Skilled nursing, wound/ostomy care, indwelling medical device care |  | Skilled nursing, wound/ostomy care, indwelling medical device care, inpatient rehabilitation |  | Skilled nursing, wound/ostomy care, indwelling medical device care, inpatient rehabilitation |  |
|  | Number of licensed beds | 200 |  | 150 |  | 120 |  |
| Residents |  | Mean | Range | Mean | Range | Mean | Range |
|  | Weekly resident census (n) | 169.8 | 157 - 179 | 79.7<sup>§</sup> | 74 - 87<sup>§</sup> | 92.4 | 88 - 99 |
|  | Weekly proportion of residents contributing fecal matter to wastewater (%) | 46.6 | 31.2 - 54.7 | 81.7<sup>§</sup> | 66.7 - 95.3<sup>§</sup> | 36.1 | 29.2 - 46.5 |
| Healthcare personnel (HCP) |  |  |  |  |  |  |  |
|  | Weekly number of HCP working at facility | 189.5 | 176 - 207 | 159.7 | 153 - 167 | 78.1 | 72 - 86 |
| Visitors |  |  |  |  |  |  |  |
|  | Weekly number of visitors recorded | 421.8 | 238 - 568 | 412.2 | 353 - 594 | 44.8 | 10 - 90 |

\*Abbreviations: SARS-CoV-2, severe acute respiratory syndrome coronavirus 2; SNF, skilled nursing facility; Min, minimum; Max, maximum; HCP, healthcare personnel
<sup>†</sup>Twenty-five (25) weekly wastewater samples and weekly epidemiologic surveys were collected from SNF A during February–July 2022, from SNF B during March–September 2022, and from SNF C during June–December 2022.
<sup>‡</sup>None of the participating skilled nursing facilities provided ventilator care.
<sup>§</sup>Resident statistics for SNF B in this table represent one wing of the facility where effluent flowed to the wastewater sampling manhole. As described in the main text of the manuscript, resident data for the entire facility was initially collected, but when it was determined that wastewater samples represented the population of just one wing, some epidemiologic data were recollected using the condensed epidemiologic data re-collection tool (Supplement 1). The remainder of data in this table was collected using the study's original weekly questionnaire (Supplement 1).

#### Plumbing Tracer Analysis and SNF B Data Re-collection

The plumbing tracer analyses performed at SNFs A and C confirmed effluent flow to the sampling manhole represented the entire resident population. At SNF B, the tracer analysis revealed that the effluent flowing to the sampling location represented only one of two wings of the facility. For full results of the tracer analyses please see Coulliette-Salmond et al. (16).

### Wastewater Results and Statistical Analyses

#### Comparison of enMF and NP Concentration Methods

There were 25 wastewater sampling events from each facility with an overall total of 75 sampling events. For each facility separately, as well as overall, the median SARS-CoV-2 RNA concentration in wastewater from enMF was higher than that from NP (SNF A: 28.4 vs. 14.9 gc/100 mL; SNF B: 85.0 vs. 60.8 gc/100 mL; SNF C: 53.0 vs. 38.6 gc/100 mL; overall: 53.0 vs. 38.6 gc/100 mL) (Figure 2). The Wilcoxson signed-rank test showed that the difference in median viral RNA concentrations was statistically significant for SNF B wastewater (p = 0.001) and overall (p = 0.001), but not for SNF A (p = 0.25) or SNF C (p = 0.20). These results indicate that, overall, enMF yielded higher SARS-CoV-2 RNA concentrations than NP. This trend was also seen with the endogenous control, *C. communis* (Supplement 4).

Based on the results of comparing the enMF and NP methods, we focused on highlighting the results from the enMF workflow which recovered overall higher virus RNA concentrations. Results presented in the next three subsections related to *SARS-CoV-2 Viral RNA Concentration in Wastewater, Sensitivity and Specificity, and Spearman’s Correlations* are from the laboratory workflow using the enMF concentration method. Statistical results from the NP concentration method, as well as assay characteristics, and results for BRSV and HCoV-OC43 using enMF and NP concentration methods, are presented in Supplement 4.

#### SARS-CoV-2 Viral RNA Concentration in Wastewater

At SNF A, SARS-CoV-2 was detected during 9/25 (36%) sampling events when accounting for quantifiable positive results and 14/25 (56%) when including DNQ results. Combined N1/N2 virus RNA concentrations ranged from 13.5 gc/100 mL to 1.51 × 10^5^ gc/100 mL (Figure 1A).

At SNF B, SARS-CoV-2 was detected during 10/25 (40%) sampling events when accounting for quantifiable positive results and 19/25 (76%) when including DNQ results. Combined N1/N2 virus RNA concentrations ranged from 25.1 gc/100 mL to 9.70 × 10^4^ gc/100 mL (Figure 1B).

At SNF C, SARS-CoV-2 was detected during 8/25 (32%) sampling events when accounting for quantifiable positive results and 15/25 (60%) when including DNQ results. Combined N1/N2 virus RNA concentrations ranged from 16.7 gc/100 mL to 5.26 × 10^5^ gc/100 mL (Figure 1C).

#### Sensitivity and Specificity

The sensitivity of using a SARS-CoV-2-positive wastewater sample to determine the presence of at least one reported SARS-CoV-2 infection among all residents was highest at SNF A (100.0%, 95% CI: 54.1 – 100.0%) and lowest at SNF B (61.5%, 95% CI: 31.6 – 86.1%; Table 2). Across all three facilities, overall sensitivity for identifying infections among all residents was 75.0% (CI: 50.9 – 91.3%; Table 2).

**Table 2:** Sensitivity and specificity of using wastewater SARS-CoV-2 concentration (using enMF concentration method^†^) to detect SARS-CoV-2 infections at three skilled nursing facilities over 25 weeks of sampling, Atlanta, Georgia, 2022.

|  | SNF A | SNF B | SNF C | Overall |
| --- | --- | --- | --- | --- |
| **All residents** |  |  |  |  |
| Sensitivity (%) | 100.0 | 61.5<sup>‡</sup> | 100.0 | 75.0 |
| Sensitivity 95% CI (%) | 54.1 – 100.0 | 31.6 -86.1<sup>‡</sup> | 2.5-100.0 | 50.9 - 91.3 |
| Specificity (%) | 84.2 | 83.3<sup>‡</sup> | 70.8 | 78.2 |
| Specificity 95% CI (%) | 60.4 - 96.6 | 51.6- 97.9<sup>‡</sup> | 48.9-87.4 | 65.0-88.2 |
| **Residents and healthcare personnel combined** |  |  |  |  |
| Sensitivity (%) | 90.0 | 56.3 | 100.0 | 74.2 |
| Sensitivity 95% CI (%) | 55.5 - 99.8 | 29.9 - 80.2 | 47.8 - 100.0 | 55.4-88.1 |
| Specificity (%) | 100.0 | 88.9 | 85.0 | 90.9 |
| Specificity 95% CI (%) | 78.2 - 100.0 | 51.8-99.7 | 62.1 - 96.8 | 78.3 - 97.5 |

\*Abbreviations: SARS-CoV-2, severe acute respiratory syndrome coronavirus 2; enMF, electronegative membrane filtration; COVID-19, coronavirus disease 2019; SNF, skilled nursing facility; CI, confidence interval
†Results presented in this table were generated with the enMF virus concentration method. Results generated with the nanoparticle virus concentration method are presented in Supplement 4, Table S4.3.
‡Data sources: SARS-CoV-2 infection count data for residents at SNF B were collected with the condensed epidemiologic data re-collection tool (Supplement 1). All other SARS-CoV-2 infection count data were obtained from the National Healthcare Safety Network (NHSN).

The specificity of using a SARS-CoV-2-negative wastewater sample to determine the absence of reported SARS-CoV-2 infections among all residents was highest at SNF A (84.2%, 95% CI: 60.4 to 96.6%) and lowest at SNF C (70.8%, 95% CI: 48.9 – 87.4%; Table 2). Across all three facilities, overall specificity for identifying infections among all residents was 78.2% (95% CI: 65.0 – 88.2%; Table 2).

Sensitivity trended slightly lower and specificity trended slightly higher when infections among all residents were combined with infections among healthcare personnel (Table 2).

#### Spearman’s Correlations

At SNF A, there were moderate to strong positive correlations between wastewater SARS-CoV-2 concentrations and SARS-CoV-2 infections among all subpopulations with Spearman’s correlation coefficient (ρ) ranging from 0.67 to 0.86 (all p-values < 0.001; Table 3). At SNF B there was a strong positive correlation among residents contributing fecal matter to wastewater ρ = 0.72 (p-value = <0.001). At SNF B, there were also moderate positive correlations among healthcare personnel (ρ = 0.48, p-value = 0.01) and among all residents and healthcare personnel combined (ρ = 0.58, p-value = 0.048, Table 3). At SNF C there were moderate positive correlations among healthcare personnel as well as all residents combined with healthcare personnel (both ρ = 0.59, p-values <0.01; Table 3).

**Table 3:**
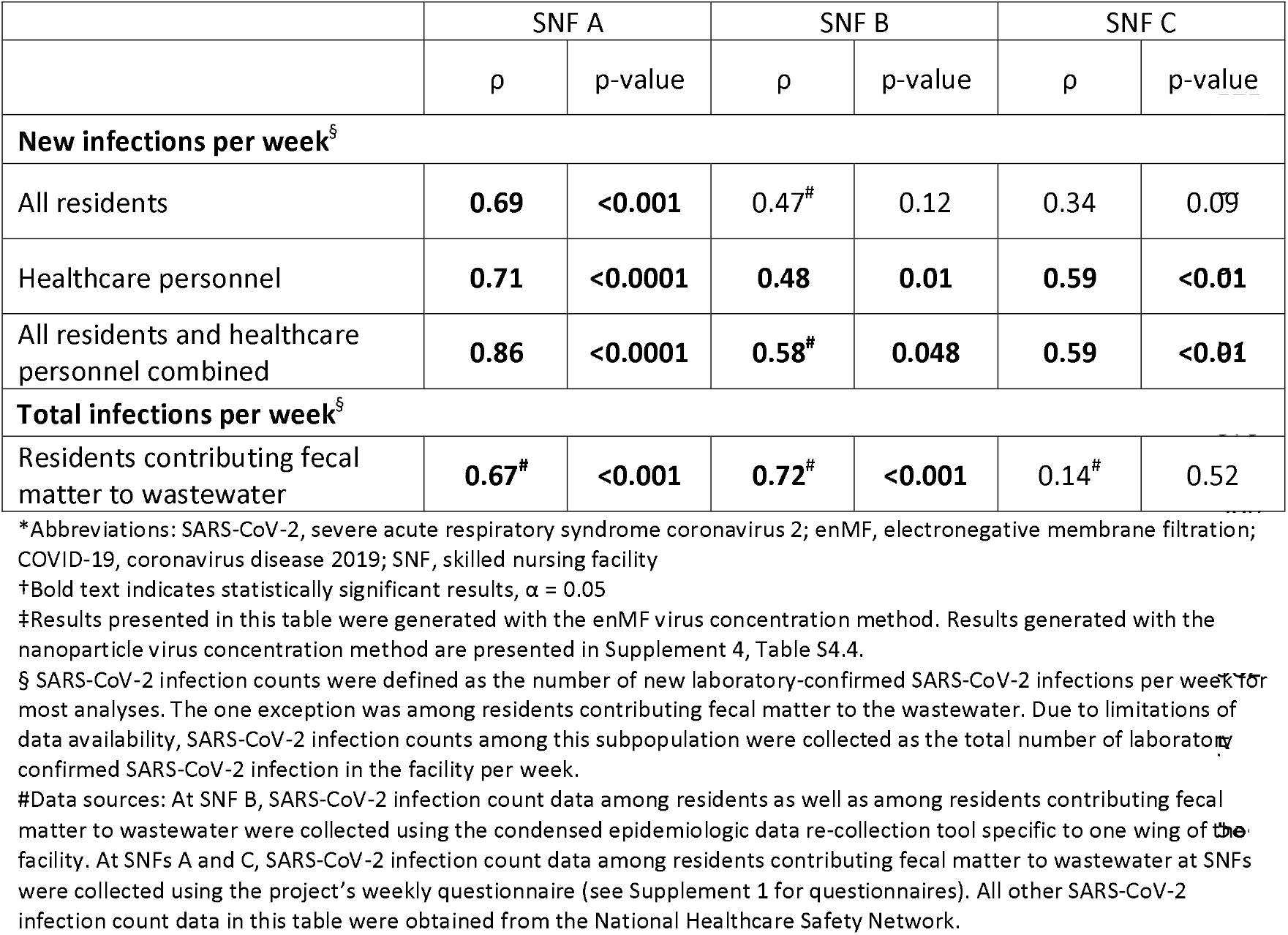
Spearman’s correlation coefficients (ρ) showing strength of correlation between wastewater SARS-CoV-2 RNA concentrations (using enMF concentration method^‡^) and SARS-CoV-2 infection counts among several subpopulations at three skilled nursing facilities over 25 weeks of sampling, Atlanta, Georgia, 2022

### Viral Sequencing

The SARS-CoV-2 lineages and their percent abundance from 11 successfully sequenced samples are presented in Table 4. Omicron lineage BA.2 was identified in one sample from May, when BA.2 was dominant across the USA (23). Lineage BA.5 was identified in the samples from June through August (n=9). Multiple BA.5 sublineages were detected in samples from the three collection sites. In four paired samples, the same dominant BA.5 sublineage was recovered by both wastewater concentration methods.

**Table 4:** SARS-CoV-2 lineages and percent abundance from genomic sequencing of wastewater samples from three skilled nursing facilities using two concentration methods, enMF and NP, Atlanta, Georgia, 2022.

| Wastewater sample date | SNF | Concentration method | SARS-CoV-2 lineages<sup>†</sup> (% abundance)<sup>§</sup> |
| --- | --- | --- | --- |
| 5/17/2022 | A | enMF | **BG.5 (98.6)**, BA.2.12.1 (1.3) |
| 6/30/2022 | C | enMF | **BA.5.5 (99.5)** |
| 7/19/2022 | A | enMF | **BA.5.1.18 (99.9)** |
| 7/26/2022 | A | NP | **BA.5.2.9 (75.7)**, BA.5.2 (20.4), BA.5.1.18 (1.7),<br>BA.5.1.23 (1.3) |
|  |  | enMF | **BA.5.2.9 (91.3)**, BA.5.1.18 (7.8) |
| 7/28/2022 | C | NP | **BF.8 (97.7)**, BA.5.2.1 (2.0) |
|  |  | enMF | **BF.8 (99.6)** |
| 8/23/2022 | B | NP | **BA.5.5 (99.6)** |
|  |  | enMF | **BA.5.5 (98.1)**, BA.5.2.20 (1.0) |
| 8/30/2022 | B | NP | **BA.5.5 (99.7)** |
|  |  | enMF | **BA.5.5 (93.6)**, BA.5 (3.0), BA.5.1.30 (3.0) |

\*Abbreviations: SARS-CoV-2, severe acute respiratory syndrome coronavirus 2; SNF, skilled nursing facility; enMF, electronegative membrane filtration; NP, nanoparticle
†Bold text indicates dominant lineage by sequencing.
‡ BG is an alias for BA.2.12.1, and BF is an alias BA.5.2.1.
§ Only lineages with ≥1 % abundance were included in this table.

## Discussion

The study presented here demonstrated moderate to strong positive correlations between wastewater SARS-CoV-2 RNA concentrations and the number of SARS-CoV-2 infections among healthcare personnel and among all residents combined with healthcare personnel at three skilled nursing facilities in the Atlanta, Georgia metropolitan area (Table 3). Strong positive correlations were also seen among a subpopulation of residents contributing fecal matter to the wastewater at SNF A and B (Table 3). The strongest correlations were found when the enMF method was used for wastewater concentration (Table 3) versus NP (Supplemental 3, Table S3.4). This approach to WWS for SARS-CoV-2 was found to be successful during a period of lower transmission during the pandemic in 2022 (24).

While only three healthcare facilities were included, our study suggests that WWS could be a valuable tool to augment SARS-CoV-2 surveillance in long-term care facilities such as SNFs. In this study, the sensitivity and specificity of using wastewater SARS-CoV-2 RNA concentration to determine the presence or absence of SARS-CoV-2 infections among residents were high at SNF A (sensitivity = 100.0%, specificity = 84.2%) and SNF C (sensitivity = 100.0%, specificity = 70.8%) where infection counts were ascertained prospectively, and moderate at SNF B (sensitivity = 61.5%, specificity = 83.3%) where infections counts from one wing of the facility needed to be re-collected retrospectively; data sources for this purpose may have been subject to recall bias.

Our study builds on two other U.S.-based studies: Gamage et al. at six Veteran Affairs nursing homes across six states and Keck et al. at eight long-term care facilities in Kentucky (9, 10). While our study had a frequency of wastewater sampling that was weekly, which was lower compared to Keck et al. (two to four times per week) and Gamage et al. (daily), in our study, sampling sensitivity among all residents was higher and specificity was similar compared to the other two studies. The presented study found that overall sensitivity among all residents was 75.0% and specificity was 78.2%; while Keck et al. found sensitivity and specificity were 48.0% and 79.9%, respectively; and Gamage et al. found sensitivity was 60.0% (specificity not reported).

When infection counts from residents and healthcare personnel were combined, this study found overall sensitivity decreased only slightly to 74.2% and specificity increased to 90.9% compared to Keck et al. where the addition of healthcare personnel reduced sensitivity to 30.6% and specificity to 79.7%. In the Keck et al. study, they mention staff worked at multiple locations within one corporation. This was not known by the research team to be the case at the three SNFs enrolled in this study and could perhaps account for the improved sensitivity and specificity when healthcare personnel were added to the analysis.

A unique aspect of this study was using two concentration methods for the duration of the study, allowing a thorough comparison of enMF and NP techniques for concentrating wastewater. Though both concentration methods yielded comparable virus recovery, we found that overall enMF produced higher SARS-CoV-2 RNA concentrations (Figure 2) and, when combined with clinical data, generally produced stronger correlations between wastewater signal and infection counts (Table 3 vs. Supplement 4, Table S4.4). Sequencing analyses demonstrated concordance across concentration methods, with both yielding the same dominant SARS-CoV-2 sublineage and thereby confirming the recovery of quality RNA from the samples. Successful WWS relies on a virus concentration method sensitive enough for low concentration pathogen detection such as is expected at a facility with resident populations in the tens to hundreds and inconsistent effluent flow.

Performing WWS at the healthcare facility level is a relatively novel tool for understanding infectious disease burden in U.S. healthcare facilities, therefore it is vital to acknowledge lessons learned while establishing a facility-level WWS study. One of the most important takeaways was that facility recruitment and selection, data collection, and communications with facilities were more resource- and time-intensive than expected and project success hinged upon the availability and knowledge of personnel at the healthcare facilities. As facility-level wastewater surveillance becomes more widespread and recognized, the time and communication burden associated with establishing such programs in healthcare settings is expected to diminish. A publication by Coulliette-Salmond et al. further discusses lessons learned and limitations while performing WWS at these three SNFs as well as from experiences with WWS partners in other states (16).

Several events impacted the trajectory of this study and affected data quality. Originally it was planned to collect wastewater samples during 25 consecutive weeks at each facility; however, high outside temperatures for staff sampling in late July through early August 2022 as well as a pause in sample collection due to essential laboratory maintenance resulted in longitudinal data gaps at SNFs B and C. At SNF C, these gaps coincided with the largest number of infections reported during the study period, causing important sampling opportunities to be missed. Additionally, plumbing tracer analyses were conducted after wastewater sampling was complete and revealed that at SNF B the sampling manhole collected effluent from only part of the facility; a limited subset of epidemiologic data was then retrospectively collected for that wing of the facility (Supplement 1). Infection count data originally collected for SNF B was from the entire facility; retrospectively recollecting these data for just one wing may have been subject to errors from limited data on patient room changes due to COVID-19 isolation protocols within the facility. It is therefore critical to perform plumbing tracer studies before facility level wastewater sampling begins to confirm the target population contributes to the effluent flowing to a wastewater access point and to ensure consistent methods of data collection across facilities. See Coulliette-Salmond et al. for a more thorough discussion of plumbing tracer studies (16).

At the time this study was conducted in 2022, facility-level WWS was not well-known to many healthcare facilities as a surveillance tool, therefore the research team dedicated time to educating the facility staff about the goals, processes, risks, and benefits of WWS for SARS-CoV-2. The research team worked with GDPH to develop a response plan that outlined roles and next steps if a wastewater result was positive. We found a response plan useful for the facility’s understanding of and participation in this WWS study. There is momentum building for facility-level WWS for additional targets, and evidence from this study and others support the utility of WWS for SARS-CoV-2 where there is interest in additional surveillance techniques and/or limited capacity for individual resident or patient screening (16, 25, 26).

## Conclusions

This study suggests that WWS could be a valuable tool to augment SARS-CoV-2 surveillance in long-term care facilities such as SNFs. Both virus concentration methods tested yielded comparable virus recovery, but we found that overall enMF produced higher SARS-CoV-2 RNA concentrations and, when combined with clinical data, generally produced stronger correlations between wastewater signal and infection counts.

## Supporting information

Supplemental_1_Questionnaires_Whitehill

Supplemental_2_WWQuality_Whitehill

Supplemental_3_dMIQE_Whitehill

Supplemental_4_AdditionalMethodsResults_Whitehill

## Data Availability

Data produced in the present study are available upon reasonable request to the authors

## Acknowledgements

We extend gratitude to Jeanne Negley and JoAnna Wagner at the Georgia Department of Public Health (GDPH) for their support and guidance reaching out to skilled nursing facilities in the Atlanta, GA metropolitan area. We also thank the staff at the skilled nursing homes enrolled in this study for their participation and dedication to this study.

## Footnote Page

^§^ See e.g., 45 C.F.R. part 46, 21 C.F.R. part 56; 42 U.S.C. §241(d); 5 U.S.C. §552a; 44 U.S.C. §3501 et seq.

## Appendices

Supplement 1: Data collection questionnaires

Supplement 2: Wastewater quality testing methods and results

Supplement 3: dMIQE checklist

Supplement 4: Additional methods and results

