## Supplemental_1_Questionnaires_Whitehill for "Insights from Wastewater Surveillance of SARS-CoV-2 in Skilled Nursing Facilities: Comparing Virus Concentration Methods for Wastewater and Correlating Wastewater Virus Concentrations with Clinical Infections, Georgia, USA, 2022"

### 2019 Novel Coronavirus Response:
#### Nursing Home Wastewater-based Epidemiology

CS357991-A 03/24/2025

### Nursing Home Wastewater Surveillance Facility Enrollment Survey

*Document purpose: To record information during the initial site visit(s) to determine the feasibility of a nursing home facility being enrolled into longitudinal study*

Name of the Facility: ______________________

Address: ______________________

County: ____________ State: ____________ Project Facility ID: ____________

CMS certification #: ____________ Staff Initials: ________ Date Assessment Completed: ________

#### Section 1. Wastewater (WW) Sampling Site Assessment

##### Facility Point of Contacts

*[CDC to enter information, gathered during calls and/or first site visit(s)]*

Administrator/Director: ____________ Director of Nursing: ____________

Infection Preventionist: ____________ Engineer/environmental/maintenance: ____________

Preferred Point of Contact (if different than the above): ____________

##### Facility Criteria for WW Study Inclusion

*[CDC to enter information, prior first site visit. Confirm during site visit(s).]*

**1.** COVID-19 case(s) within the last 90 days* ☐ Yes ☐ No

**2.** Facility in EIP catchment ☐ Yes ☐ No

**3.** On-going/admission MDRO screening ☐ Yes ☐ No

**4.** Current # resident census >75 ☐ Yes ☐ No

***If yes to #1 or 2, see the Nursing Home Facility Information, Section 2, Question #11***

**5.** If there are no current COVID-19 cases within the facility and SARS-CoV-2 is detected in the facility wastewater, we would suggest a Point Prevalence Survey be conducted for residents and staff to help mitigate spread. Is this something your facility would be open to? ☐ Yes ☐ No

##### Facility, Grounds, and Engineering Questions

*[CDC to enter information during site visit(s)]*

**1.** Manhole size (diameter, thickness) and other characteristics:

______________________

a. Distance from facility (feet): ________

b. Energy source/outlet available for autosampler? ☐ Yes ☐ No

c. Sufficient room for sampling equipment and barricade? ☐ Yes ☐ No

d. Description of manhole location (e.g., external influences, low point where flooding occurs, near the entrance or parking lot)

______________________________

**2.** Cleanout (this information provides a backup location to sample if needed or appropriate)

a. Location: ______________________________

b. Description: ______________________________

**3.** Potable Water Meter Location:

______________________________

**4.** Facility amenable to equipment being set-up long-term? ☐ Yes ☐ No

**5.** Facility engineering and/or grounds staff available daily if issues arise? ☐ Yes ☐ No

**6.** Does the facility plumbing system have any chlorine injectors installed to treat wastewater before it enters the county sewer system? ☐ Yes ☐ No

***If yes, specify area installed:***

______________________________

**7.** Communication: Is facility open to a sign on the wastewater sampling equipment and a handout to the public if they have questions? ☐ Yes ☐ No

#### Safety Questions

*[CDC to enter information during site visit(s)]*

**1.** Adequate parking and set-up area for CDC staff? ☐ Yes ☐ No

**2.** Is sampling location in a safe area away from vehicle traffic (e.g., parking lot, driveway, fire lane)? ☐ Yes ☐ No

***If yes, describe:***

______________________________

**3.** Is the sampling location in a safe area, away from foot traffic (e.g., entrance, visitor walkway, resident patio)? (check and describe): ☐ Yes ☐ No

**4.** Assess the sampling location for any safety concerns, describe below:

______________________________

#### Section 2. Nursing Home Facility Information

##### Facility Enrollment Questions

*[CDC to request information from facility during calls, share via email to facility, facility enters the information, and CDC obtains during first site visit]*

**The following questions are meant to gather information about the size of your facility, the types of residents for which your facility provides care, and other people that may contribute to wastewater.**

**1.** What types of care (also described as patient acuity) does your facility provide? *Select all that apply:*

☐ Skilled nursing
☐ Wound/ostomy care
☐ Inpatient rehabilitation
☐ Ventilator care
☐ Tracheostomy care
☐ Indwelling medical devices (e.g., central vascular catheter, urinary catheter)

**2.** Are there other care populations or healthcare facilities on premises (i.e., vSNF, co-located wards, assisted living, LTACH, freestanding inpatient rehabilitation facility)? ☐ Yes ☐ No

***If yes, please describe:***

______________________________

***If yes, does sampled manhole receive wastewater from these facilities?***

______________________________

**3.** Does the facility systematically track visitors, and can the number of visitors per week be shared with CDC? ☐ Yes ☐ No

***This helps us to estimate the number of people who may be contributing to the facility's wastewater.***

**4.** Current resident census or occupied beds: ___________

**5.** Please answer the following about vaccination status in the table below:

| Persons | Vaccination status | Answer |
|---|---|---|
| **Residents** | Number of fully vaccinated[1] residents | |
| | Number of partially vaccinated[2] residents | |
| | Number of unvaccinated[3] residents | |
| | Number of boosted[4] residents | |
| | Name of vaccine(s) used | |
| **Healthcare personnel (HCP)** | Total number of HCP | |
| | Number of fully vaccinated[1] HCP | |
| | Number of unvaccinated[3] HCP | |
| | Number of boosted[4] HCP | |
| | Name of vaccine(s) used | |

[1] **Fully Vaccinated:** Individuals who have completed a two-dose mRNA series (Pfizer, Moderna) or a single dose vaccine series (Janssen) AND are ≥ 14 days from vaccine series completion.

[2] **Partially Vaccinated:** Individuals who have received at least one dose of a two-dose vaccine series or one dose of a single-dose series but do not yet meet criteria for being fully vaccinated.

[3] **Unvaccinated:** Individual has not received any doses of any COVID-19 vaccine

[4] **Boosted:** Individual with completed COVID-19 vaccination receiving booster/additional doses

6. Do you have any COVID-19 testing requirements for those who are newly admitted, readmitted, or that regularly leave the facility? ☐ Yes ☐ No

7. (For MDROs only) Are any residents screened on admission (or otherwise) for antibiotic-resistant organisms? ☐ Yes ☐ No

______________________________________________

8. In the absence of an active outbreak do you perform any regular testing of residents other than the situations described above?

______________________________________________

9. What is your current routine testing protocol and frequency for employees? Note: this is testing that occurs regardless of an ongoing outbreak.

______________________________________________

10. How does the facility identify cases among HCP? *(Select all that apply)*

☐ **Symptom-based strategy** (i.e., symptom screens prior to each shift)
☐ **Test-based strategy** (i.e., point prevalence surveys)
☐ Antigen test, *please specify*:______________________________
☐ RT-PCR
☐ Both
☐ **Other**, *please specify*: ______________________________

11. What is the minimum number of days a HCP must stay-at-home before returning to work after a COVID-19 positive test?

__________

12. What is your facility's testing protocol in response to a newly identified case of SARS-CoV-2 infection of a resident/staff? Please check all that apply:

☐ Perform testing of all residents and staff (regardless of whether those being tested had contact with the newly identified case)
☐ Perform contact tracing and test only those residents/staff who had contact with the new case
☐ Perform testing of all residents and staff on the unit(s) where the newly identified staff member worked/resident stayed prior to testing positive
☐ Other (please describe): ______________________________

13. How often are residents and/or staff tested during an ongoing (active) outbreak?

☐ Weekly
☐ Twice weekly
☐ Monthly
☐ Other (please describe): ______________________________

14. Has the facility experienced a COVID-19 case/outbreak in the last 60 days? ☐ Yes ☐ No

a. Date index case tested positive: __________

b. Was the first case a HCP, resident, or visitor?______________________________

c. If a resident, did this resident test positive within 7 days of facility admission? ☐ Yes ☐ No

d. If a HCP, did this person work during the 48-hrs leading up to the date of SARS-CoV-2 detection? ☐ Yes ☐ No

e. How was the first COVID-19 case identified? (fell ill then tested, case was symptomatic and then tested, or identified through screening?)

______________________________

f. How many of the current residents were present during the outbreak? ______________________________

g. When was the most recent case of COVID-19 ? ______________________________

h. Was this case a HCP, resident, or visitor? ______________________________

i. Total number of cases in this outbreak: **HCP:** __________ **Residents:** __________

**15.** Have you performed facility-wide testing (i.e., point prevalence surveys (PPS) to test all residents in your facility for SARS-CoV-2), in the past 60 days? Testing may be for any reason (e.g., outbreak response, surveillance). ☐ Yes ☐ No

***If yes, please list the dates and numbers in the table below:***

| PPS Date | Total residents tested | Total test-positive resident cases | Total HCP tested | Total test-positive HCP cases |
|---|---|---|---|---|

#### **Section 3.** Fecal Waste Management and Environmental Cleaning

*Since our study is focused on detection of SARS-CoV-2 from fecal material in wastewater, we have a few questions related to how your facility handles fecal waste among residents. We would also like to collect information on environmental cleaning and disinfecting products, as these may also impact the ability to detect SARS-CoV-2.*

**1.** What is the average number of residents continually diapered in the facility? ______________________________

**2.** How do staff members typically dispose feces from the following containers?

| Fecal waste container | Disposed of in toilet? |
|---|---|
| Bedpan | ☐ Yes ☐ No |
| Bedside commode | ☐ Yes ☐ No |
| Diaper/Bed pad | ☐ Yes ☐ No |

**3.** If possible, we would like to review all cleaning and disinfecting products used (including wipes, sprays, laundry, and dish detergents/solutions).

***Note to CDC staff:***
***Would recommend taking photos of product labels whenever possible, ask to see the environmental cleaning cart or closet.***

#### Environmental Cleaner #1:

Name: ______________________________

Intended use (as reported by facility): ______________________________

Method of disposal (e.g., poured down drain; thrown in trash): ______________________________

#### Environmental Cleaner #2:

Name: ______________________________

Intended use (as reported by facility): ______________________________

Method of disposal (e.g., poured down drain; thrown in trash): ______________________________

#### Environmental Cleaner #3:

Name: ______________________________

Intended use (as reported by facility): ______________________________

Method of disposal (e.g., poured down drain; thrown in trash): ______________________________

#### Environmental Cleaner #4:

Name: ______________________________

Intended use (as reported by facility): ______________________________

Method of disposal (e.g., poured down drain; thrown in trash): ______________________________

#### Environmental Cleaner #5:

Name: ______________________________

Intended use (as reported by facility): ______________________________

Method of disposal (e.g., poured down drain; thrown in trash): ______________________________

#### Section 4. Additional documents to collect

*[CDC to request during calls and/or during the site visit(s)]*

- **Facility map:** Please provide a facility map with number of beds per room indicated.
- **Kitchen, laundry, cleaning schedule:** Please provide any routine schedules that would list detergents, disinfectants, and large volumes of water being introduced into the wastewater system. (e.g., note volumes if a liquid disinfectant is used, percentage/strength, expiration)
  - Timing of commercial dish washer use
  - Onsite laundry and timing
  - Dump unused remaining disinfectants down the drain in the morning (e.g., unused bleach from the day before)

### Nursing Home Wastewater Surveillance Weekly Questionnaire

*Document purpose: To record weekly information during sampling events*

#### **Section 1.** Wastewater Sampling Questionnaire [Outside satellite person]

*[CDC to enter information during site visit for sample collection]*

**1.** CDC staff conducting the questionnaire: ______________________ Date Completed: __________

**2. Facility Point of contact Information:**

Name(s): ______________________

Contact Information: ______________________

Date/Time of contact attempt(s): ______________________

**3.** Has anyone asked questions to the CDC staff or the front desk, whether from the public, residents, and/or facility staff? ☐ Yes ☐ No

***If yes, add here:***

______________________

#### **Section 2.** Nursing Home Facility Information [Epi-assessment]

*[CDC to ask facility point of contact and enter information during site visit for sample collection]*

**1.** Document number of visitors per day since the last facility visit (N/A if visitation not occurring for current week):

☐ N/A

| Date | # of Visitors | Notes |
|---|---|---|

**2.** Please answer the following about the cumulative vaccination status in the table below:

| Persons | Sub category | Number or percent |
|---|---|---|
| **Residents** | Total number of residents (current census): | |
| | Number of fully vaccinated[1] residents | |
| | Number of partially vaccinated[2] residents | |
| | Number of unvaccinated[3] residents | |
| | Number of boosted[4] residents | |
| **Healthcare personnel (HCP)** | Number of HCP | |
| | Number of fully vaccinated[1] HCP | |
| | Number of partially vaccinated[2] HCP | |
| | Number of unvaccinated[3] HCP | |
| | Number of boosted[4] HCP | |

[1] **Fully Vaccinated:** Individuals who have completed a two-dose mRNA series (Pfizer, Moderna) or a single dose vaccine series (Janssen) AND are ≥ 14 days from vaccine series completion.

[2] **Partially Vaccinated:** Individuals who have received at least one dose of a two-dose vaccine series or one dose of a single-dose series but do not yet meet criteria for being fully vaccinated.

[3] **Unvaccinated:** Individual has not received any doses of any COVID-19 vaccine.

[4] **Boosted:** Individual with completed COVID-19 vaccination receiving booster/additional doses

#### **Section 3.** Fecal Waste Management [Epi-assessment]

*[Since our study is focused on detection of SARS-CoV-2 from fecal material in wastewater, we have a few questions related to how your facility handles fecal waste among residents.]*

**1.** Were there instances of toilets or other plumbing clogging that impacted the facility since the last questionnaire (**Date here:** __________ ) ☐ Yes ☐ No

**2.** Document number of residents able to use the toilet:

| Residents able to use the toilet | # of residents |
|---|---|
| Active infection (i.e., resident in COVID unit) | |
| Recovered, within 30 days of infection | |
| Other residents in facility using the toilet | |
| Potential SC2 Fecal Contribution | |

**3.** Were there any events in the last 7 days where a large volume of disinfectant was disposed of down the drain? ☐ Yes ☐ No

***If yes, describe:***

a. What day and time? __________

b. What was the disinfectant/chemical used? __________

c. Disinfectant volume: __________

d. Disinfectant concentration: __________

#### **Section 4.** Positive Case Survey [Epi-assessment]

***IF there was a positive case, please complete the following questions.***

**1.** Has the facility experienced a COVID-19 case and/or outbreak since date of last survey? ☐ Yes ☐ No

**Date here:** __________

a. Date index case tested positive: __________

b. Was the first case a resident? ☐ Yes ☐ No

c. If a resident, did this resident test positive within 7 days of facility admission? ☐ Yes ☐ No

d. If a HCP, did this person work during the 48-hrs leading up to the date of SARS-CoV-2 detection? ☐ Yes ☐ No

e. How was this first case of COVID-19 case identified? (e.g., fell ill then tested, case was symptomatic and then tested, identified through screening)

__________

f. When was the most recent case of COVID-19? __________

g. Was this case a HCP, resident, or visitor? __________

h. Total number of cases in this outbreak: **HCP:** ________ **Residents:** ________ **Visitors:** ________

**2.** Additional notes or observations potentially impacting sampling (i.e., manhole integrity, weather/flooding, building work, foot traffic, etc.):

__________

### Condensed Epidemiological Data Re-Collection Tool
#### Skilled Nursing Facility B

**Sampling week date range – Date:** ________________

| | |
|---|---|
| **1.** What was the total number of residents in the ____________ wing* each week? | **Number:** ____________ |
| **2.** Among residents using the toilet in the ____________ wing, how many had an **active SARS-CoV-2 infection** each week (e.g., resident in COVID unit)? | **Number:** ____________ |
| **3.** Among residents using the toilet in the ____________ wing, how many had **recovered from SARS-CoV-2 infection in the previous 30 days**? | **Number:** ____________ |
| **4.** How many **other** residents in the ____________ wing were able to use the toilet each week? | **Number:** ____________ |
| **5. Total residents in the ____________ wing potentially contributing to SARS-CoV-2 in fecal** matter each week (sum of questions 2 thru 4)? | **Number:** ____________ |
| **6.** Did the facility experienced a COVID-19 case and/or outbreak in the ____________ wing during this week? | ☐ Yes ☐ No |
| **7.** If there was a COVID-19 outbreak in the ____________ wing during this timeframe, what was the date the index case tested positive? | **Date:** ______________ |
| **8.** When was the most recent case of COVID-19 in the ____________ wing during each week? | **Date:** ______________ |
| **9.** What was the total number of residents in the ____________ wing who were ill during the outbreak? | **Number:** ____________ |
| **10.** What was the total number of healthcare personnel working in the ____________ wing who were ill during the outbreak? | **Number:** ____________ |

*At SNF B, a plumbing tracer analysis revealed that the effluent from only one of two wings of the facility flowed to the manhole sampling location.
