## Supplemental_2_WWQuality_Whitehill for "Insights from Wastewater Surveillance of SARS-CoV-2 in Skilled Nursing Facilities: Comparing Virus Concentration Methods for Wastewater and Correlating Wastewater Virus Concentrations with Clinical Infections, Georgia, USA, 2022"

Supplement 2: Wastewater Sample Quality Testing

### 1. **Wastewater Quality Testing**

The wastewater quality testing methods, including *Escherichia coli* and total coliform analysis, were conducted following the approach detailed in the pilot study, Santiago et al., 2025 (1). Table S2.1 summarizes the wastewater quality indicators across three skilled nursing facilities (SNF), showing the median values, range, and interquartile ranges (IQR) for each parameter. The number of samples (n) varies across both facilities and parameters due to several data quality considerations including suspected disinfection events, inconclusive or anomalous results, autosampler malfunctions, and incomplete documentation. To address these data quality challenges, we used imputation and data exclusion criteria. Exclusions were used for samples with inconclusive results, if we suspected a large volume of disinfectants was added to the wastewater stream, or insufficient supportive metadata. Imputations were applied only when missing values could be reliably estimated, such as when measurements were below detection limits or instances where instrument readings were recorded but not quantifiable. These steps were implemented to ensure the reliability and comparability of the dataset while minimizing bias in downstream analyses (see Table S2.1 footnotes for details).

| **Table S2.1:** Descriptive statistics of wastewater quality indicators from skilled nursing facilities (SNF) A, B, and C in Atlanta, Georgia, 2022 | | | | | | | | | | | | |
| --- | --- | --- | --- | --- | --- | --- | --- | --- | --- | --- | --- | --- |
|  | **SNF A** | | | | **SNF B** | | | | **SNF C** | | | |
| **Wastewater quality indicator** | **No.**  **(n)** | **Median** | **Range** | **IQR** | **No.**  **(n)**^¶^ | **Median** | **Range** | **IQR** | **No.**  **(n)** | **Median** | **Range** | **IQR** |
| Temperature (⁰C) | 25 | 25.6 | 14.2 | 3.1 | 26 | 25.8 | 16.1 | 2.2 | 25 | 25.6 | 19.0 | 8.3 |
| pH | 25 | 10.6 | 4.9 | 2.1 | 26 | 7.5 | 5.8 | 2.7 | 25 | 8.4 | 5.1 | 2.1 |
| Electrical conductivity (µS/cm) | 25 | 436.5 | 1035.5 | 317.0 | 26 | 265.9 | 450.4 | 209.4 | 25 | 268.3 | 1530.0 | 139.8 |
| Total dissolved solids (mg/L) | 25 | 349.0 | 504.0 | 184.0 | 25^#^ | 211 | 350 | 76.0 | 25 | 196.0 | 2467.0 | 138.0 |
| Total suspended solids (mg/L) | 25 | 118.0 | 566.5 | 138.8 | 25^#^ | 51.5 | 238.5 | 43.0 | 25 | 65.0 | 481.5 | 71.5 |
| Biochemical oxygen demand (mg/L) | 25^†^ | 153.0 | 411.5 | 100.7 | 25^#^ | 163 | 351 | 97.0 | 25 | 106.0 | 658.7 | 91.3 |
| Total organic carbon (mg/L) | 25 | 98.8 | 347.7 | 99.1 | 25^#^ | 71.2 | 143.0 | 80.2 | 25 | 69.9 | 343.9 | 46.4 |
| Total organic nitrogen (mg/L) | 25 | 116.0 | 97.37 | 14.2 | 25^#^ | 8.5 | 25.8 | 7.84 | 24^††^ | 16.7 | 66.2 | 15.6 |
| Total coliforms (log_10_ MPN/100ml) | 24^‡^ | 5.5 | 7.3 | 6.1 | 26 | 5.3 | 7.1 | 6.0 | 25 | 7.0 | 1.9 | 0.3 |
| *E. coli* (log_10_ MPN/100ml) | 23^§^ | 4.9 | 6.3 | 5.2 | 26^**^ | 4.7 | 6.7 | 5.2 | 25 | 6.2 | 1.7 | 0.5 |
| *Abbreviations: No., number of samples; IQR, interquartile range; MPN, most probable number  ^†^A value below the detection limit was imputed for biochemical oxygen demand at SNF A to ensure consistency in statistical analysis.  ^‡^No coliforms were detected at SNF A for one sampling date. It is suspected a large volume of disinfectant was likely introduced into the wastewater stream and no adequate explanation was provided by the facility. This data point was excluded.  ^§^No *E. coli* were detected at SNF A for two sampling dates. It is suspected a large volume of disinfectant was likely introduced into the wastewater stream and no adequate explanation was provided by the facility. These data points were excluded.  ^¶^SNF B had 26 total samples collected and tested for water quality and SARS-CoV-2. One sample had an invalid PCR result for SARS-CoV-2 and an additional sample was collected and tested for SARS-CoV-2 and wastewater quality.  ^#^Data for total dissolved solids, total suspended solids, biochemical oxygen demand, total organic carbon, and total organic nitrogen at SNF B were unavailable for one sampling event due to an autosampler malfunction for grab sample collection.  ^**^During one sampling event at SNF B, *E. coli* was detected but not quantifiable; an imputed value was used.  ^††^A total organic nitrogen value was imputed for one sampling event at SNF C due to concentrations falling below the detection limit. | | | | | | | | | | | | |

### **2. Spearman’s Rank Coefficient Correlation Analysis of Wastewater Quality Parameters**

Given the non-parametric nature of the data, Spearman’s rank correlation was used to assess the relationships between wastewater quality parameters, *Escherichia coli,* total coliform bacteria, and severe acute respiratory syndrome coronavirus 2 (SARS-CoV-2) concentrations across the three facilities. The correlograms in Figure S2.1 panels A – C represent these correlations, with circle size indicating the strength of the association and color representing the direction of the correlation, where yellow-to-orange hues denote positive correlations and blue-green hues denote negative correlations. Only statistically significant correlations (p < 0.05) are visualized in Figure S2.1.

#### **Figure S2.1:** Spearman correlation (ρ) matrices of wastewater quality indicators and severe acute respiratory syndrome coronavirus 2 (SARS-CoV-2) concentrations produced with two virus concentration methods, nanoparticle and electronegative membrane filtration, from skilled nursing facility (SNF) A, B, and C in Atlanta, GA, 2022.

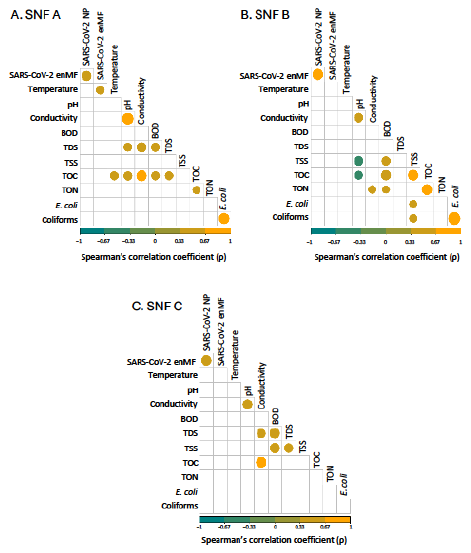

Footnotes Figure S2.1:

Abbreviations: NP, nanoparticle concentration method; enMF, electronegative membrane filtration concentration method; BOD, biochemical oxygen demand; TDS, total dissolved solids; TSS, total suspended solids; TOC, total organic carbon; TON, total organic nitrogen
