## Supplemental_3_dMIQE_Whitehill for "Insights from Wastewater Surveillance of SARS-CoV-2 in Skilled Nursing Facilities: Comparing Virus Concentration Methods for Wastewater and Correlating Wastewater Virus Concentrations with Clinical Infections, Georgia, USA, 2022"

Supplement 3: Checklist completed for this study providing Minimum Information for Publication of Quantitative Digital PCR Experiments (dMIQE)(1)

| ITEM TO CHECK | PROVIDED Y/N | COMMENT |
| --- | --- | --- |
| 1. SPECIMEN |  |  |
| Detailed description of specimen type and numbers | Y | METHODS |
| Sampling procedure (including time to storage) | Y | METHODS |
| Sample aliquotation, storage conditions and duration | Y | METHODS |
| 2. NUCLEIC ACID EXTRACTION |  |  |
| Description of extraction method including amount of sample processed | Y | METHODS |
| Volume of solvent used to elute/resuspend extract | Y | METHODS |
| Number of extraction replicates | Y | METHODS |
| Extraction blanks included? | Y | METHODS |
| 3. NUCLEIC ACID ASSESSMENT AND STORAGE |  |  |
| Method to evaluate quality of nucleic acids | Y | METHODS |
| Method to evaluate quantity of nucleic acids (including molecular weight and calculations when using mass) | N | Not performed |
| Storage conditions: temperature, concentration, duration, buffer, aliquots | Y | METHODS |
| Clear description of dilution steps used to prepare working DNA solution | Y | METHODS |
| 4. NUCLEIC ACID MODIFICATION |  |  |
| Template modification (digestion, sonication, pre-amplification, bisulphite, etc.) | N | Not applicable |
| Details of repurification following modification if performed | N | Not applicable |
| 5. REVERSE TRANSCRIPTION |  |  |
| cDNA priming method and concentration | N | Not applicable |
| One or two step protocol (include reaction details for two step) | Y | METHODS |
| Amount of RNA added per reaction | Y | METHODS |
| Detailed reaction components and conditions | Y | METHODS |
| Estimated copies measured with and without addition of RT^*^ | Y | METHODS |
| Manufacturer of reagents used with catalogue and lot numbers | Y | METHODS |
| Storage of cDNA: temperature, concentration, duration, buffer and aliquots | Y | METHODS |
| 6. dPCR OLIGONUCLEOTIDES DESIGN AND TARGET INFORMATION |  |  |
| Sequence accession number or official gene symbol | N | NC_045512.2; N gene (2) |
| Method (software) used for design and *in silico* verification | N | N/A |
| Location of amplicon | N | N gene region; positions ~28,000–29,000 bp (CDC Panel) |
| Amplicon length | Y | METHODS |
| Primer and probe sequences (or amplicon context sequence)^†^ | Y | METHODS (3, 4) |
| Location and identity of any modifications | N | Not applicable |
| Manufacturer of oligonucleotides | Y | METHODS |
| 7. dPCR PROTOCOL |  |  |
| Manufacturer of dPCR instrument and instrument model | Y | METHODS |
| Buffer/kit manufacturer with catalogue and lot number | Y | METHODS |
| Primer and probe concentration | Y | METHODS |
| Pre-reaction volume and composition (incl. amount of template and if restriction enzyme added) | Y | METHODS |
| Template treatment (initial heating or chemical denaturation) | N | Not applicable |
| Polymerase identity and concentration, Mg++ and dNTP concentrations^‡^ | N | Not applicable commercial kit (Cat # 186-3023/3024) (5) |
| Complete thermocycling parameters | Y | METHODS |
| 8. ASSAY VALIDATION |  |  |
| Details of optimization performed | N | Not applicable validated by manufacturer |
| Analytical specificity (vs. related sequences) and limit of blank (LOB) | Y | METHODS |
| Analytical sensitivity/LoD and how this was evaluated | Y | METHODS |
| Testing for inhibitors (from biological matrix/extraction) | Y | METHODS |
| 9. DATA ANALYSIS |  |  |
| Description of dPCR experimental design | Y | METHODS |
| Comprehensive details negative and positive of controls (whether applied for QC or for estimation of error) | Y | METHODS |
| Partition classification method (thresholding) | Y | METHODS |
| Examples of positive and negative experimental results (including fluorescence plots in supplemental material) | N | Not applicable |
| Description of technical replication | Y | METHODS |
| Repeatability (intra-experiment variation) | Y | METHODS |
| Reproducibility (inter-experiment/user/lab etc. variation) | Y | METHODS |
| Number of partitions measured (average and standard deviation) | N | Not performed |
| Partition volume | N | Not performed |
| Copies per partition (λ or equivalent) (average and standard deviation) | N | Not performed |
| dPCR analysis program (source, version) | Y | METHODS |
| Description of normalization method | N | Not applicable |
| Statistical methods used for analysis | Y | METHODS |
| Data transparency | raw data available on request | raw data available on request |
| *Assessing the absence of DNA using a no RT assay (or where RT has been inactivated) is essential when first extracting RNA. Once the sample has been validated as DNA-free, inclusion of a no-RT control is desirable, but no longer essential.  †Disclosure of the primer and probe sequence is highly desirable and strongly encouraged. However, since not all commercial pre-designed assay vendors provide this information when it is not available assay context sequences must be submitted (Bustin et al. Primer sequence disclosure: A clarification of the miqe guidelines. Clin Chem 2011;57:919-21.)  ‡Details of reaction components is highly desirable, however not always possible for commercial disclosure reasons. Inclusion of catalogue number is essential where component reagent details are not available. | | |

5.	Bio-Rad Laboratories. ddPCR Supermix for Probes (No dUTP).Catalog No. 186-3023/3024; <https://www.bio-rad.com/sites/default/files/webroot/web/pdf/lsr/literature/10026868.pdf>.
