## Supplemental_4_AdditionalMethodsResults_Whitehill for "Insights from Wastewater Surveillance of SARS-CoV-2 in Skilled Nursing Facilities: Comparing Virus Concentration Methods for Wastewater and Correlating Wastewater Virus Concentrations with Clinical Infections, Georgia, USA, 2022"

Supplement 4: Additional Methods and Results

### 1. **Droplet Digital PCR Assay Results**

| **Table S4.1:** Wastewater sample result determination using droplet count from droplet digital PCR (ddPCR) assay | | | |
| --- | --- | --- | --- |
| Result | Total droplets | Positive SARS-CoV-2 droplets | Number of triplicates meeting droplet criteria in column 3 |
| Positive | >10,000 | ≥3 | 3 of 3, 2 of 3 |
| DNQ | >10,000 | 1 or 2 | 3 of 3, 2 of 3, 1 of 3 |
| DNQ | >10,000 | ≥3 | 1 of 3 |
| Negative | >10,000 | 0 | 3 of 3 |
| Inconclusive^†^ | <10,000 | N/A | N/A |
| *Abbreviations: PCR, polymerase chain reaction; SARS-CoV-2, severe acute respiratory syndrome coronavirus 2; DNQ, detected but not quantifiable; N/A, not applicable.  †Recommend repeating assay at least once more before determining inconclusive. | | | |

### 2. **Vaccination Rates**

| **Table S4.2:** Weekly SARS-CoV-2 vaccination rates among residents and healthcare personnel at three skilled nursing facilities over 25 weeks^†^, Atlanta, GA, 2022 | | | | | | | |
| --- | --- | --- | --- | --- | --- | --- | --- |
|  | | **SNF A** | | **SNF B** | | **SNF C** | |
| **Residents** | | **Mean** | **Range** | **Mean** | **Range** | **Mean** | **Range** |
|  | Weekly resident census (n) | 169.8 | 157 - 179 | 118.5 | 112-124 | 92.4 | 88 - 99 |
|  | Weekly proportion of residents fully vaccinated^‡^ against SARS-CoV-2 infection (%) | 91.6 | 88.5 - 97.7 | 98.3 | 97.5- 98.4 | 89.3 | 80.9 - 91.2 |
|  | Weekly proportion of residents partially vaccinated^§^ against SARS-CoV-2 infection (%) | 0.9 | 0 - 4.0 | 0.2 | 0 – 0.9 | 0.9 | 0 - 3.3 |
| **Healthcare personnel (HCP)** | |  | | | | | |
|  | Weekly number of HCP working at facility (n) | 189.5 | 176 - 207 | 159.7 | 153 - 167 | 78.1 | 72 - 86 |
|  | Weekly proportion of HCP fully vaccinated^‡^ against SARS-CoV-2 infection (%) | 96.7 | 95.5 - 100.0 | 99.4 | 99.3 - 99.4 | 98.2 | 96.1 - 98.8 |
|  | Weekly proportion of HCP partially vaccinated^§^ against SARS-CoV-2 infection (%) | 0.9 | 0 - 3.0 | 0 | 0 - 0 | 0.6 | 0 - 2.6 |
| * Abbreviations: SARS-CoV-2, severe acute respiratory syndrome coronavirus 2; SNF, skilled nursing facility; HCP, healthcare personnel  † Twenty-five (25) weekly epidemiologic surveys were collected from SNF A during February–July 2022, from SNF B during March–September 2022, and from SNF C during June–December 2022.  ‡ Fully vaccinated: Individuals who have completed a two-dose mRNA series (Pfizer, Moderna) or a single dose vaccine series (Janssen) AND are ≥ 14 days from vaccine series completion.  § Partially vaccinated: Individuals who have received at least one dose of a two-dose vaccine series or one dose of a single-dose series but do not yet meet criteria for being fully vaccinated. | | | | | | | |

### 3. **Nanoparticle Virus Concentration Results**

At skilled nursing facility (SNF) A, SARS-CoV-2 virus RNA was detected during 5/25 (20%) sampling events when accounting for quantifiable positive results and 12/25 (48%) when including detected but not quantifiable (DNQ) results; viral RNA concentrations ranged from 11.9 gc/100 mL to 3.2 X 10^4^ gc/100 mL (Figure S4.1A). At SNF B, SARS-CoV-2 viral RNA was detected during 3/25 (12%) sampling events when accounting for quantifiable positive results and 13/25 (52%) when including DNQ results; viral RNA concentrations ranged from 34.4 gc/100 mL to 5.23 x 10^4^ gc/100 mL (Figure S4.1B). At SNF C, SARS-CoV-2 viral RNA was detected during 6/25 (24%) sampling events when accounting for quantifiable positive results and 11/25 (64%) when including DNQ results; viral RNA concentrations ranged from 34.4 gc/100 mL to 7.96 x 10^4^ gc/100 mL (Figure S4.1C).

For the Nanoparticle (NP) concentration method, sensitivity of using a SARS-COV-2 positive wastewater sample to determine the presence of at least one SARS-CoV-2 infection among all residents was highest at SNF A (100.0 %, 95% CI: 39.7 to 100.0%) and lowest at SNF B (25.0%, 95% CI 5.5 to 57.2%). Overall sensitivity among all residents was 47.1% (95% CI: 23.0 to 72.2%) across all three facilities. The specificity of using a SARS-CoV-2 negative wastewater sample to determine the absence of SARS-CoV-2 infection among all residents was highest at SNF B (100.0%, 95% CI: 75.3 to 100.0%) and lowest at SNF C (79.2 %, 95% CI: 57.9 to 92.9%). Overall, specificity among all residents was 89.7% (95% CI: 78.8 to 96.1%) across all three facilities (Table S4.3).

At SNF A, there were moderate positive correlations between wastewater SARS-CoV-2 viral RNA concentrations and SARS-CoV-2 infections among all subpopulations with Spearman’s correlation coefficient (ρ) ranging from 0.43 to 0.54 (all p-values <0.05). At SNF B, there was a strong positive correlation between wastewater SARS-CoV-2 viral RNA concentrations and SARS-CoV-2 infections among all residents combined with healthcare personnel (ρ = 0.74, p-value < 0.01) and moderate positive correlations among all residents (ρ = 0.68, p-value = 0.02), among healthcare personnel (ρ = 0.40, p-value = 0.049), and among residents contributing fecal matter to wastewater(ρ = 0.56, p-value = 0.02). At SNF C, there were no significant correlations between wastewater SARS-CoV-2 viral RNA concentrations and SARS-CoV-2 infections among all subpopulations (Table S4.4).

#### **Figure S4.1:** Wastewater SARS-CoV-2 viral RNA concentrations (using the nanoparticle virus concentration method^†^) and SARS-CoV-2 infection counts among residents and healthcare professionals over 25 weeks at three skilled nursing facilities (panels A – C), Atlanta, GA, 2022

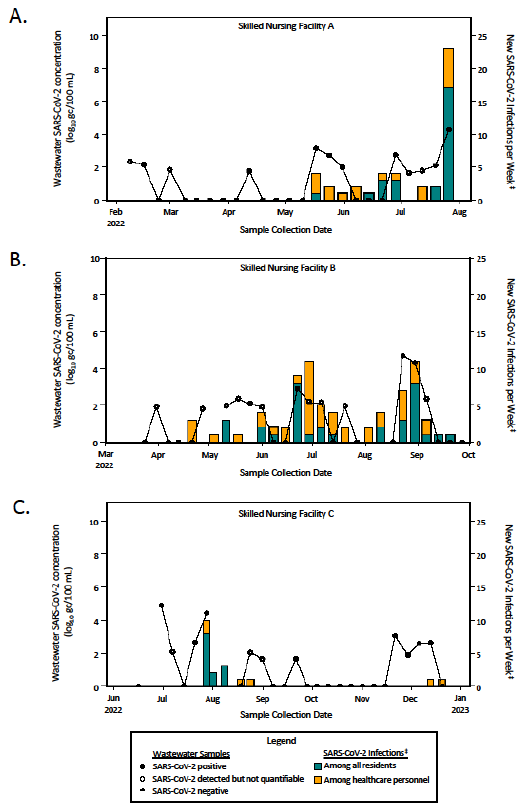

Figure S4.1 Footnotes:

*Abbreviations: SARS-CoV-2, severe acute respiratory syndrome coronavirus 2; gc, genome copies

†Results presented in this figure were generated with the nanoparticle concentration method. Results generated with the electronegative membrane filtration concentration method are presented in the main manuscript, Figure 1.

‡Data sources for SARS-CoV-2 infections: counts among healthcare personnel as well as among all residents at skilled nursing facilities (SNFs) A and C were obtained from the National Healthcare Safety Network Long-Term Care Facility COVID-19 Report; infection counts among residents in one wing of at SNF B were obtained from the condensed epidemiologic data re-collection tool (Supplement 1).

| **Table S4.3:** Sensitivity and specificity of using wastewater SARS-CoV-2 concentration (using nanoparticle concentration method^†^) to detect SARS-CoV-2 infections at three skilled nursing facilities over 25 weeks of sampling, Atlanta, Georgia, 2022 | | | | |
| --- | --- | --- | --- | --- |
|  | SNF A | SNF B | SNF C | Overall |
| **All residents** | | | | |
| Sensitivity (%) | 100.0 | 25.0^‡^ | 100.0 | 47.1 |
| Sensitivity 95% CI (%) | 39.7 - 100.0 | 5.5 - 57.2 | 2.5 - 100.0 | 23.0 - 72.2 |
| Specificity (%) | 95.2 | 100.0^‡^ | 79.2 | 89.7 |
| Specificity 95% CI (%) | 76.9 - 99.9 | 75.3 - 100^‡^ | 57.9 - 92.9 | 78.8 - 96.1 |
| **All residents and healthcare personnel combined** | | | | |
| Sensitivity (%) | 50.0 | 18.8 | 40.0 | 32.3 |
| Sensitivity 95% CI (%) | 18.7 - 81.3 | 4.1 - 45.65 | 5.3 - 85.3 | 16.7 - 51.4 |
| Specificity (%) | 100.0 | 100.0 | 80.0 | 90.9 |
| Specificity 95% CI (%) | 78.2 - 100.0 | 66.4 – 100.0 | 56.3 - 94.3 | 78.3 - 97.5 |
| *Abbreviations: SARS-CoV-2, severe acute respiratory syndrome coronavirus 2; SNF, skilled nursing facility; CI, confidence interval  †Results presented in this table were generated with the nanoparticle concentration method. Results generated with the electronegative membrane filtration virus concentration method are presented in the main manuscript, Table 2.  ‡Data sources: SARS-CoV-2 infection count data for residents at SNF B were collected with the condensed epidemiologic data re-collection tool (Supplement 1). All other SARS-CoV-2 infection count data were obtained from the National Healthcare Safety Network (NHSN). | | | | |

| **Table S4.4:** Spearman’s correlation coefficients (ρ) showing strength of correlation between wastewater SARS-CoV-2 viral RNA concentration (using nanoparticle concentration method^‡^) and SARS-CoV-2 infection counts among several subpopulations at three skilled nursing facilities over 25 weeks of sampling, Atlanta, Georgia, 2022 | | | | | | |
| --- | --- | --- | --- | --- | --- | --- |
|  | SNF A | | SNF B | | SNF C | |
|  | ρ | p-value | ρ | p-value | ρ | p-value |
| **New infections per week^§^** | | | | | | |
| All residents | **0.43** | **0.03** | **0.68^#^** | **0.02** | 0.32 | 0.12 |
| Healthcare personnel | **0.50** | **0.01** | **0.40** | **0.049** | 0.23 | 0.26 |
| All residents and healthcare providers combined | **0.54** | **<0.01** | **0.74**^#^ | **<0.01** | 0.23 | 0.26 |
| **Total infections per week^§^** | | | | | | |
| Residents contributing fecal matter to wastewater | **0.50**^#^ | **0.01** | **0.56^#^** | **0.02** | 0.34^#^ | 0.10 |
| *Abbreviations: SARS-CoV-2, severe acute respiratory syndrome coronavirus 2; SNF, skilled nursing facility  †Bold text indicated statistically significant results, α = 0.05  ‡Results presented in this table were generated with the NP concentration method. Results generated with the electronegative membrane filtration concentration method are presented in the main manuscript, Table 3.  § SARS-CoV-2 infection counts were defined as the number of new laboratory-confirmed SARS-CoV-2 infection per week for most analyses. The one exception was among residents contributing fecal matter to the wastewater. Due to limitations of data availability, SARS-CoV-2 infection counts among this subpopulation were collected as the total number of laboratory-confirmed SARS-CoV-2 infections in the facility per week.  #Data sources: At SNF B, SARS-CoV-2 infection count data among residents and among residents contributing fecal matter to wastewater at were collected using the condensed epidemiologic data re-collection tool specific to one wing of the facility. At SNFs A and C, SARS-CoV-2 infection count data among residents contributing fecal matter to wastewater were collected using the project’s weekly questionnaire (see Supplement 1 for questionnaires). All other SARS-CoV-2 infection count data in this table were collected from the National Healthcare Safety Network. | | | | | | |

### **2. *Carjivirus communis*, HCoV-OC43, and BRSV Positive Controls Results**

Median concentration (n = 75 across three facilities) of *Carjivirus communis* was 6.9 log_10_ gc/100 mL (min-max: 2.4 to 9.8 log_10_ gc/100mL) for results produced using the enMF virus concentration method and 5.5 log_10_ gc/100 mL (min-max: 4.2 to 6.8 log_10_ gc/100 ml) for those produced using the NP method. A paired non-parametric Wilcoxson signed-rank test indicated that there is significant difference in median *C. communis* detection between the concentration method groups (p-value = 1.32 x 10^-9^).

At each of the three facilities (n= 25 per facility), the enMF method consistently detected higher *C. communis* viral concentrations compared to the NP method, with statistically significant differences as indicated by a paired Wilcoxon signed-ranked test. The median concentrations for enMF vs. NP were 7.0 vs. 5.1 log_10_ gc/100 mL at SNF A (p= 1.669 x 10^-5^), 6.9 vs. 5.7 log_10_ gc/100 mL at SNF B (p= 0.0003), and 6.9 vs. 5.7 log_10_ gc/100 mL at SNF C (p= 0.02). (Figure S4.2).

A pilot study by Santiago et al. determined median (range) percent recoveries of the extraction control, human coronavirus OC43 (HCoV OC43), by concentration method: 1.31% (0.00%-3.68%) and 1.50% (0.66%-2.73%) for enMF and NP (n=23 samples each) (1). For this study, HCoV-OC43 extraction control was spiked in multiple wastewater samples from each facility (n = 9 from SNF A, n = 10 from SNF B, n = 6 from SNF C) as spot checks of the percent recoveries throughout the course of the study and revealed similar recoveries; see table S4.5 for HCoV-OC43 recovery statistics. Other published wastewater studies were not found to report extract control recovery data, thus not allowing for a broader comparison.

Bovine respiratory syncytial virus (BRSV) process control was spiked in every wastewater sample (n=25 per facility). The median percent recoveries were 88.62% (0.96%-503.42%) using enMF for concentration and 65.03% (13.08%-200.42%) using NP for concentration; see table S4.6 for detailed BRSV recovery statistics. Compared to the pilot study, Santiago et al. 2025, this study demonstrated a higher median percent recovery using enMF, and similar recovery results using NP (enMF: 69.84% [0.00%-948.75%]; NP 13.02% [1.27%-70.86%]). Beyond Santiago et al., there were no additional peer-reviewed publications for a direct comparison of process control percent recoveries. However, there are similar processing approaches in the literature, Othman et al. 2023 and Farkas et al. 2024, that have recovery data for SARS-CoV-2 for which BRSV is a surrogate virus (2, 3). Additionally, one study used murine hepatitis virus to determine processing recovery percentages, another accepted surrogate for SARS-CoV-2 (4). For SARS-CoV-2, Othman et al. reported mean percent recovery of 25.59 ±5.04% using enMF, while Farkas et al. demonstrated a median percent recovery of 7.70% using NP (2, 3). This study had higher percent recoveries for both methods using BRSV. Additionally, this study also had higher percent recovery using enMF as compared to the study using murine hepatitis virus that found a mean of 65.7% (±23.8%) (4).

#### **Figure S4.2:** Concentration of *Carjivirus communis* viral RNA detected by droplet digital PCR at three skilled nursing facilities using two virus concentration methods, electronegative membrane filtration (enMF) and Nanotrap® magnetic virus particles (NP)

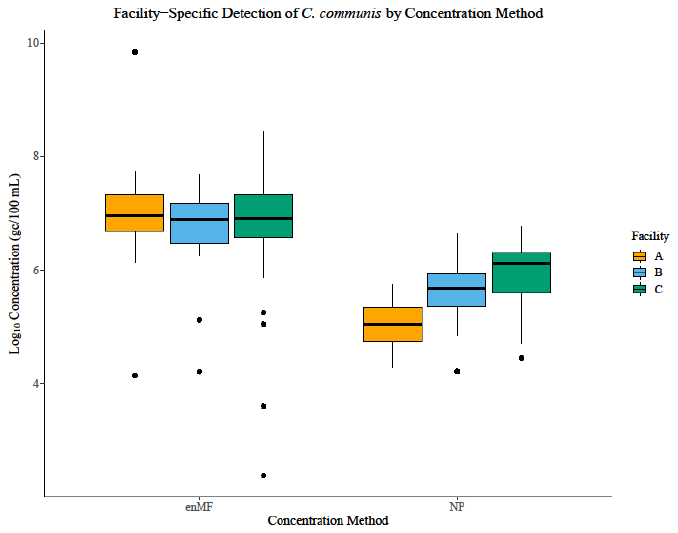

Figure S4.2 Footnotes:

Abbreviations: gc, genome copies; SNF, skilled nursing facility; enMF, electronegative membrane filtration; NP, nanoparticle

Box-and-whisker plots represent the median, interquartile range, and range of *C. communis* viral RNA concentration. Individual data points outside the whiskers are considered outliers. Outliers are defined as values greater than 1.5 times the IQR above the third quartile (Q3) or less than 1.5 times the IQR below the first quartile (Q1).

| **Table S4.5:** Percent recovery of extraction control human coronavirus-OC43 using two virus concentration methods, electronegative membrane filtration (enMF) vs. nanoparticle (NP), across skilled nursing facilities A, B, and C in Atlanta, GA, 2022 | | | | |
| --- | --- | --- | --- | --- |
|  | SNF A | SNF B | SNF C | All facilities combined |
| Total number of samples spiked (n) | 9 | 10 | 6 | 25 |
| Total positive samples (n) from spiked samples (%) | 9 (100) | 10 (100) | 6 (100) | 25 (100) |
| enMF | | | | |
| Median concentration (gc/ul) | 8.98 x 10^4^ | 1.16 x 10^5^ | 2.12 x 10^5^ | 1.16 x 10^5^ |
| Median % recovery | 1.44 | 1.63 | 2.87 | 1.63 |
| Minimum % recovery | 0.10 | 0.05 | 0.01 | 0.05 |
| Maximum % recovery | 2.69 | 35.87 | 3.63 | 3.63 |
| NP | | | | |
| Median concentration (gc/ul) | 7.30 x 10^4^ | 1.38 x 10^5^ | 1.54 x 10^5^ | 1.38 x 10^5^ |
| Median % recovery | 1.49 | 2.23 | 2.21 | 2.21 |
| Minimum % recovery | 0.42 | 0.03 | 1.22 | 0.42 |
| Maximum % recovery | 16.04 | 17.98 | 2.81 | 16.04 |

| **Table S4.6:** Percent recovery of process control bovine respiratory syncytial virus RNA using two virus concentration methods, electronegative membrane filtration (enMF) vs. nanoparticle (NP), across skilled nursing facilities A, B, and C in Atlanta, GA, 2022 | | | | |
| --- | --- | --- | --- | --- |
|  | SNF A | SNF B | SNF C | All facilities combined |
| Total number of samples spiked (n) | 25 | 25 | 25 | 75 |
| Total positive samples (n) from spiked samples (%) | 25 (100) | 25 (100) | 25 (100) | 75 (100) |
| enMF | | | | |
| Median concentration (gc/µl) | 3.5 x 10^1^ | 4.25 x 10^1^ | 1.47 x 10^1^ | 3.50 x 10^1^ |
| Median % recovery | 88.62 | 140.84 | 55.55 | 88.62 |
| Minimum % recovery | 1.48 | 0.96 | 0.01 | 0.96 |
| Maximum % recovery | 503.42 | 332.31 | 1269.43 | 503.42 |
| NP | | | | |
| Median concentration (gc/µl) | 4.71 x 10^0^ | 3.63 x 10^1^ | 2.21 x 10^1^ | 2.21 x 10^1^ |
| Median % recovery | 13.25 | 107.84 | 65.03 | 65.03 |
| Minimum % recovery | 1.58 | 17.78 | 13.08 | 13.08 |
| Maximum % recovery | 107.40 | 200.42 | 204.33 | 200.42 |

### **3. SARS-CoV-2 Virus Sequencing from Wastewater Samples**

#### Methods

SARS-CoV-2-positive samples were selected for sequencing based on real-time PCR cycle quantification (Cq) values <35. Viral RNA was amplified using random hexamers to generate cDNA, followed by a targeted SARS-CoV-2 multiplex PCR amplification protocol as described previously (5). Sequencing was performed using the Illumina DNA Library Prep kit strategy (Cat # 20018704, 20018705) on a MiSeq instrument (Illumina Inc., San Diego, CA, USA). Additional sequencing was performed using the Nanopore GridION Sequencer as confirmation of sequence availability. Sequences were analyzed using the CFSAN Wastewater Analysis Pipeline with Freyja (version 1.3.11) to identify the mixture of SARS-CoV-2 lineages in the specimen (6). The Freyja analysis was performed separately for the R1 and R2 paired read sets from each sample, which identified the same lineages at similar proportions in both read sets. Genome coverage was measured by mapping the SARS-CoV-2 genome GenBank accession no. NC_045512.2 (Wuhan-Hu-1 isolate) with IRMA to produce consensus sequences (7). However, due to the pooled nature of each specimen—potentially containing genetic material from multiple infections—lineages were not assigned to consensus genomes. Historical SARS-CoV-2 lineage frequencies were obtained from the weekly weighted dataset described by Ma *et al*. and published to data.cdc.gov on January 6, 2023 (8).

The available genome sections were compared with the known locations for determining a lineage as acquired from COV-Spectrum by aligning to the Wuhan-Hu-1 genome sequence, and missing regions were assessed for their importance to a lineage call (9, 10).

#### Results

A total of 18 concentrated wastewater samples from three nursing homes in the Atlanta, Georgia area were selected for sequencing. Sixteen of these samples were accepted for sequencing based on a viral load threshold of Cq<35 by real-time PCR (11). Greater than 60% genome coverage was achieved for 11 of the 16 samples. Lower genome coverage in the remaining samples suggests the presence of SARS-CoV-2, albeit with limited or degraded template material. The population of lineages within each sample was estimated by analyzing the reads using Freyja (12). Greater lineage diversity was observed in specimens with lower percentage of genome coverage, likely reflecting a technical artifact rather than true biological variation. This artifact arises because lineage-defining mutations are concentrated in the S gene region, not randomly dispersed across the genome (13). As a result, pooled genome populations lacking S-gene sequences may have imprecise lineage proportion estimates.
